# Compound heterozygous mutations in the kinase domain of IKKα lead to immunodeficiency and immune dysregulation

**DOI:** 10.1101/2024.05.17.24307356

**Authors:** Quentin Riller, Boris Sorin, Charline Courteille, Duong Ho-Nhat, Tom Le Voyer, Jean-Christophe Debray, Marie-Claude Stolzenberg, Olivier Pellé, Thomas Becquard, María Rodrigo Riestra, Laureline Berteloot, Mélanie Migaud, Laure Delage, Marie Jeanpierre, Charlotte Boussard, Camille Brunaud, Aude Magérus, Victor Michel, Camille Roux, Capucine Picard, Cécile Masson, Christine Bole-Feysot, Nicolas Cagnard, Aurélien Corneau, Isabelle Meyts, Véronique Baud, Jean-Laurent Casanova, Alain Fischer, Emmanuel Dejardin, Anne Puel, Cécile Boulanger, Bénédicte Neven, Frédéric Rieux-Laucat

## Abstract

IKKα, encoded by *CHUK*, is crucial in the non-canonical NF-κB pathway and part of the IKK complex activating the canonical pathway alongside IKKβ. Absence of IKKα cause fetal encasement syndrome in human, fatal in utero, while an impaired IKKα-NIK interaction was reported in a single patient and cause combined immunodeficiency. Here, we describe compound heterozygous variants in the kinase domain of IKKα in a female patient with hypogammaglobulinemia, recurrent lung infections, and Hay-Wells syndrome-like features. We showed that both variants were loss-of-function. Non-canonical NF-κB activation was profoundly diminished in stromal and immune cells while the canonical pathway was partially impaired. Reintroducing wild-type *CHUK* restored non-canonical NF-κB activation. The patient had neutralizing autoantibodies against type I IFN, akin to non-canonical NF-κB pathway deficiencies. Thus, this is the first case of bi-allelic *CHUK* mutations disrupting IKKα kinase function, broadening non-canonical NF-κB defect understanding and suggesting IKKα’s role in canonical NF-κB target gene expression in human.

## Introduction

The NF-κB pathway is divided into the canonical (or classical) and the non-canonical (or alternative) pathways. They both lead *in fine* to the translocation in the nucleus of homo or hetero dimers formed by the particular association of 5 transcription factors including RelA (also called p65), c-Rel, RelB, NF-κB1 (the precursor p105 of p50) and NF-κB2 (the precursor p100 of p52) *(Smale, 2012)*. These dimers are sequestered in the cytoplasm by inhibitors of NF-κB (IκB family) such as the IκΒα inhibitor for the canonical pathway or the unprocessed form of *NFKB2* p100 for the non-canonical one *(Zandi et al., 1997; Baeuerle and Baltimore, 1988b; a, 1989)*. The canonical pathway provides a rapid response to the activation of various receptors (Toll-like receptors, Tumor Necrosis Factor Receptor 1, Ectodysplasin A receptor, B-cell receptor and T-cell receptor) and leads to the transcription of multiple pro-inflammatory and pro-survival genes *(Schnappauf and Aksentijevich, 2020; Baltimore, 2009)*. The “IKK complex” containing the catalytic subunits IKKα and IKKβ and a regulatory subunit IKKγ (NEMO)(Zandi et al., 1997; Mercurio et al., 1997; Ea et al., 2006; Israël, 2010) is essential to this process. Once activated, the effector serine/threonine kinases of the IKK complex phosphorylates the IκΒα inhibitor on two serines (and other IκΒ members such as IκΒε or IκΒβ), leading to their polyubiquitination and degradation by the *26S* proteasome within seconds. Free from inhibition, canonical dimers containing p65 or c-Rel can translocate to the nucleus where they exert their transcriptional activities. In contrast, the non-canonical NF-κB pathway is fully activated after hours of receptor engagement. These receptors are all members of the TNFR superfamily and includes for example CD40, LTβR and BAFF-R, all of which have been implicated in the development of lymphoid organs, normal maturation and activation of B and T cells. The non-canonical pathway is dependent on IKKα *(Dejardin et al., 2002)*. Before receptor activation, the activating kinase of IKKα called NIK is constantly degraded by a cIAP1-2/TRAF2/TRAF3 complex *(Sun, 2017; Bista et al., 2010)*. After ligation of receptors, TRAF3 is degraded, NIK accumulates and phosphorylates IKKα at serine 176/180 *(Dejardin et al., 2002)*. Once activated, NIK and IKKα phosphorylate NF-κB2 (p100), tagging it for polyubiquitination *(Dejardin et al., 2002; Senftleben et al., 2001)*. The partial cleavage by the proteasome of p100 into p52 results in RelB:p52 dimers translocation to the nucleus and transcription of target genes. Depending on the engaged receptor, the non-canonical pathway regulates the development of secondary lymphoid organs and their organization (via LTβ receptor), the medullary thymic epithelial cell differentiation and AIRE expression (via RANK), thymocyte emigration (LTβ receptor), B-cell survival, development, and maturation (via CD40 and BAFF receptor) and finally controls the bone formation and homeostasis (via RANK) *(Sun, 2017, 2012)*. While murine models have revealed the relative importance of each component of the NF-κB signaling pathways, the study of patients with inborn errors of immunity and deleterious mutations in NF-κB transcription factors or signaling components reveals their non-redundant roles and their implication in humans *(Bista et al., 2010; Hu et al., 2001; Li et al., 1999; Takeda et al., 1999; Franzoso et al., 1998; Tucker et al., 2007; Yin et al., 2001; Bousfiha et al., 2020; Tangye et al., 2022; Zhang et al., 2017, 30; Barnabei et al., 2021)*. Regarding the non-canonical pathway, bi-allelic NIK deficiency as well as complete RELB deficiency has been associated to combined immune deficiency with immune dysregulation *(Willmann et al., 2014; Merico et al., 2015; Ovadia et al., 2017; Sharfe et al., 2015)*. Heterozygous *NFKB2* variants have been reported with various mechanisms from complete deficiency in both p100 and the processed p52 to uncleavable p100 leading to p52 loss-of-function (LOF) and p100 gain-of-function (GOF) phenotype *(Le Voyer et al., 2023; Chen et al., 2013)*. Patients with heterozygous *NFKB2* variants exhibit a wide variety of symptoms due to immune dysregulation associated with various levels of immunodeficiency from hypogammaglobulinemia to combined immunodeficiency. Until recently, no living patients were described with *CHUK* (coding IKKα) deleterious mutations. Homozygous nonsense null *CHUK* mutations led to the fetal encasement syndrome that is lethal *in utero* similarly to mice knock-out for this gene who died quickly after birth *(Lahtela et al., 2010)*. Of note, parents carrying heterozygous nonsense variants were reported to be healthy, suggesting tolerance to haploinsufficiency for IKKα *(Lahtela et al., 2010)*. Recently, a single patient presenting with a combined immunodeficiency and immune dysregulation was reported carrying a homozygous missense mutation in *CHUK* (p. Y580C). This mutation led to the selective impairment of the non-canonical NF-κB pathway by impairing the ability of IKKα to interact with its activator NIK *(Bainter et al., 2021)*.

In this report, we describe the case of a patient who suffered from syndromic immunodeficiency and various immune dysregulation due to an autosomal recessive form of IKKα kinase deficiency.

## Results

### A patient with immunodeficiency, lymphoma, and compound heterozygous missense mutations in *CHUK*

The patient was first reported in a clinical description by Khandelwal et al and was found associated with a heterozygous mutation. *(Khandelwal et al., 2017)*. Briefly the patient (P) was the third child of an otherwise healthy non-consanguineous kindred (**Figure 1A**). The patient’s presentation was reminiscent of Hay-Wells syndrome with Pierre Robin sequence, ectodermal dysplasia, blepharophimosis, alopecia, and abnormal dental enamel *(Cole et al., 2009; Celli et al., 1999)*. She also had skeletal malformations of feet and hands requiring surgery in infancy. In addition, she had a profound hypogammaglobulinemia associated with recurrent otitis and pneumonia requiring immunoglobulin replacement therapy from the first year of life. She also experienced chronic onychomycosis of the finger nails. A diagnosis of celiac disease was made based on villous atrophy, following the exploration of growth failure without growth hormone deficiency. This villous atrophy did not respond well to gluten-free regimen and it was hypothesized to be a consequence of a chronic enteric viral infection or immune dysregulation and T-cell infiltration rather than a *bona fide* celiac disease *(Riller et al., 2023; Strohmeier et al., 2022; Klocperk et al., 2022)*. Peripheral blood immunophenotyping showed a lack of memory B-cells and an excess of naïve T cells as well as an excess of effector memory CD8+ T cells and a CD4+ lymphocytosis (**Supplementary Table 1**). Biological follow-up showed a fluctuant cytolytic hepatitis with reactional unspecific hepatitis on liver biopsies and mild sinusoidal fibrosis. As a teenager, she was finally diagnosed with a non-GC, EBER-, diffuse large B-cell lymphoma (DLBCL) involving the lungs, associated with pleural effusion (**Figure 1B**). Immunohistochemistry was consistent with non-germinal center (non-GC) DLBCL. The patient died of a massive pulmonary hemorrhage few days after the second infusion of chemotherapy. Sanger sequencing of *TP63,* the known gene to be mutated in Hay-Wells syndrome showed no mutation and a CGH array did not show any abnormality *(Khandelwal et al., 2017)*. The initial description reported a *de novo* heterozygous missense variant in *CHUK* (encoding IKKα) leading to the substitution of a histidine into arginine at position 142 of the amino-acid sequence, close to the catalytic domain of the kinase domain of IKKα (*CHUK* c. 425A>G, p.H142R, **Figure 1C**). However, we sequenced the *CHUK cDNA* from fibroblasts and peripheral blood mononuclear cells (PBMCs) and we identified a second missense variant (CHUK c.569 C>T, **Figure 1C**) leading to the substitution of a proline into a leucine at position 190 (p.P190L) within the kinase domain of IKKα. Sanger sequencing of the proband’s SV40-fibroblasts’s and activated T cells’ genomic DNA and SV40-fibroblasts complementary DNA (cDNA) confirmed both heterozygous variants (**Supplementary Figure 1A**). Both variants were absent from public databases (gnomAD v4) and were predicted deleterious by *in silico* scores with a CADD score of 25.1 and 30 respectively, well above the MSC threshold of 3.13. (**Figure 1E**) *(Zhang et al., 2018; Itan et al., 2016)*. This patient was thus carrying two germline predicted deleterious variants in the kinase domain of IKKα.

**Figure 1.**
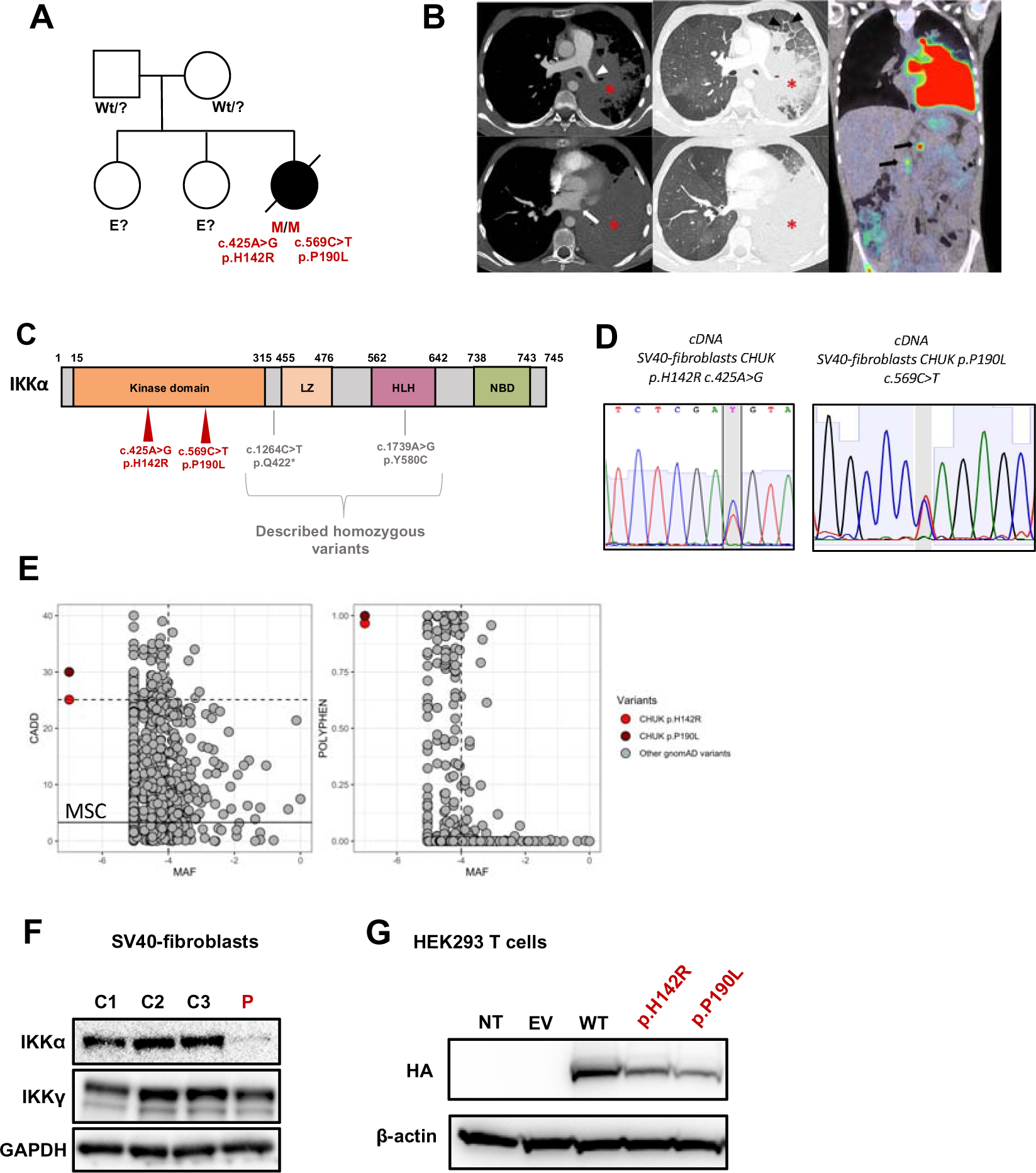
Pedigree, clinical manifestations, and mutations of a patient with immunodeficiency. **A**. Pedigree of the family; “wt” indicates a wt genotype for *CHUK* **B**. Aspect of primary pulmonary lymphoma on the CT scan: Consolidation of the lower two thirds of the left lung (asterisk) with invasion of the mediastinum, infiltration of the left pulmonary artery (white arrowhead) and complete thrombosis of the left pulmonary veins (white arrow). Thickening of the interlobular septa in the remainder of the ventilated lung, consistent with venous stasis or carcinomatous lymphangitis (black arrowhead). No normal lymph nodes or adenomegaly were individualized in the mediastinum or in the pulmonary hilum. A few small lymph nodes were visible in the axillary areas. The PET-CDFDG 18F performed within 48 hours showed hypermetabolism of the lesions and in three coeliomesenteric lymphadenopathies (black arrows) **C**. Localisation of the mutation on the protein sequence of IKKα. The patient mutations are shown in red while previously reported variants are in gray. **D**. Sanger sequencing of RT-PCR products (cDNA) from the indicated cells. **E**. Combined annotation dependent depletion score versus minor allele frequency (CADD-MAF) (left panel) and polyphen score versus MAF (right panel). The patient variants are in red and dark red and the other gnomAD variants in gray. MSC: mean significance cutoff for *CHUK* CADD score (3.13). **F**. Protein expression of IKKα and IKKγ οn protein lysates from healthy controls SV40-fibroblasts and the patient (P, in red). GAPDH was used a protein loading control. **G**. Protein expression of HA-tagged IKKα on protein lysates from transfected HEK293T cells with the different variants, the wt (WT) or an empty vector (EV). The patient variants are displayed in red.

Given that the parents’ DNAs were not available, we performed a TOPO-TA cloning of a RT-PCR product encompassing the two mutations on cDNA from patient’s SV40-fibroblasts. The sequencing of colonies showed that the mutations were bi-allelic (**Supplementary Figure 1B**). Therefore, we concluded that the patient was compound heterozygous for these predicted deleterious variants in *CHUK*. No quantitative difference was found by RTqPCR of *CHUK* mRNA (**Supplementary Figure 1C**) in fibroblasts suggesting that both alleles are equally expressed. Conversely, total IKKα protein expression was lower in SV40-fibroblasts than in the controls suggesting a potential instability of the mutated proteins (**Figure 1F**). The coding sequence of *CHUK* was cloned into a plasmid containing an N-terminal HA-tag sequence and site-directed mutagenesis was performed to obtain the patient’s variants *(Bainter et al., 2021)*. Transfection experiments of these variants or the WT in HEK293T cells showed a slightly decreased expression for both variants as compared to the wild-type thereby confirming that the IKKα p.H142R and p.P190L mutants could be less stable than the wild-type IKKα (**Figure 1G**).

### Perturbed immune phenotype in peripheral blood mononuclear cells

We analyzed the immune phenotype from thawed Peripheral Blood Mononuclear Cells (PBMCs) sampled and cryopreserved during the 5 years before the patient’s death, using cytometry by time of flight (CyTOF). Unsupervised clustering allowed the identification of 35 immune subsets (**Figure 2A, Supplementary Figure 2A-B**). On this single time point analysis, we observed a profound naïve and memory B cells lymphopenia (**Figure 2B**). In addition, plasmablasts were absent and a subclustering of B cells revealed an excess of CD19^+^CD20^+^CD27^-^IgD^+^CD38^high^ consistent with transitional B cells, a result in agreement with previously described immune phenotype in the context of non-canonical NF-κB defects (**Supplementary Figure 3A**) *(Willmann et al., 2014; Schlechter et al., 2017; Le Voyer et al., 2023; Chen et al., 2013; Bainter et al., 2021)*. The T cell compartment was characterized by a relative excess of naïve CD4+ and CD8+ T cells as previously observed in patients with a defective canonical NF-κB pathway (**Figure 2B, Supplementary Figure 2B**). This patient had an excess of highly activated HLA-DR+ CD38^high^ CD127^low^ effector memory CD8+ T cells, a cluster that we and others have already described to be associated to chronic viral infections (**Supplementary Figure 2B**) *(Riller et al., 2023; Klocperk et al., 2022; Paiardini et al., 2005)*. In contrast, we observed a decrease of Th2 and Th17 polarized memory CD4+ T cells, in accordance with the involvement of IKKα in the generation of Th17 in the recently reported bi-allelic loss-of-function *CHUK* variant p.Y580C (**Supplementary Figure 3B**) *(Bainter et al., 2021)*.

**Figure 2.**
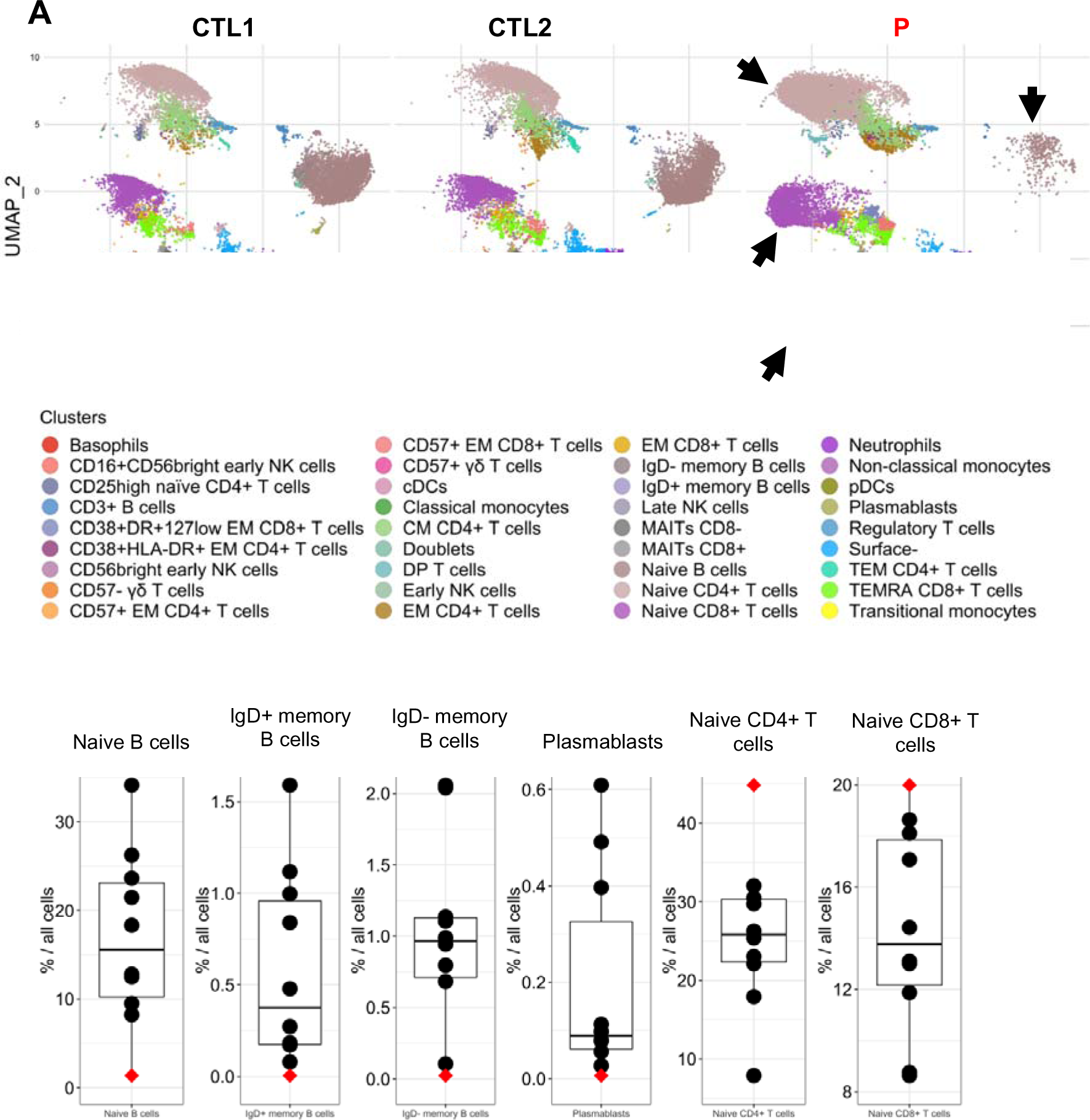
Mass-cytometry analysis of peripheral blood mononuclear cells from the patient and controls. **A**. Uniform Manifold Approximation and Projection (UMAP) of patient cells (P) and 2 representative controls (CTL1 and CTL2) with colors corresponding to the identified clusters (below the UMAPs). Arrows are showing clusters with obvious different proportions compare to controls on the patient UMAP **B**. Proportion of some identified immune subsets among all PBMCs or among memory CD4+ T cells (patient in red, controls in black), as indicated.

### Both IKKα variants were unable to activate non-canonical NF-κB *in vitro* and are loss-of-function for kinase activity

To specifically assess the non-canonical NF-κB pathway activity in an ectopic expression model without interference with the canonical one, we generated a HEK293T RELA^-/-^ cell line in which we co-expressed IKKα, RELB and NFKB2 in addition to a κB-dependent firefly luciferase expression plasmid and a thymidine kinase-dependent renilla luciferase plasmid (**Figure 3A**). By transfecting increasing amounts of IKKα WT we observed a dose-dependent increase of the relative luciferase intensity, showing that this model was relevant to assess IKKα activity in the non-canonical NF-κB pathway (**Figure 3B**). The transfection of IKKα p.H142R or IKKα p.P190L failed to induce any NF-κB activity, as compared to the empty vector condition as well as p.Q422*, a known IKKα null variant *(Lahtela et al., 2010)*. Given that the luciferase intensity was equal to the empty vector condition, this experiment suggested a pure loss-of-function for both *CHUK* variants (**Figure 3B**). Of note the expression of the IKKα p.Y580C variant, known to have a preserved kinase activity but an impaired binding to NIK *(Bainter et al., 2021)*, led to a luciferase activity comparable to that of the wt IKKα. These results indicated that this experimental setting was NIK independent and allowed to evaluate the intrinsic kinase activity of IKKα, for which both studied variants were at least hypomorphic.

**Figure 3.**
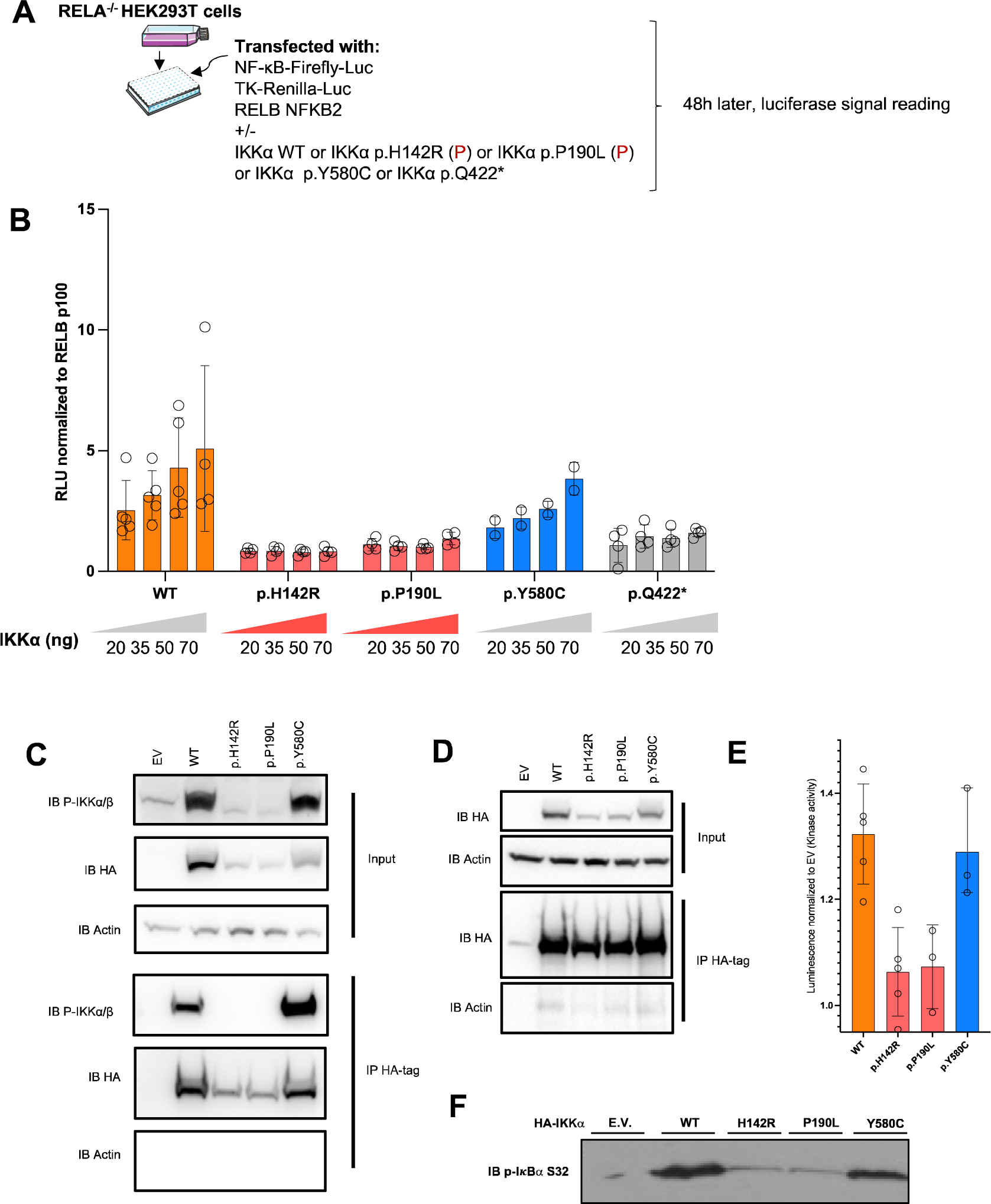
Non-canonical NF-κB activity in an ectopic expression system and kinase assays. **A**. Schematic description of the experiment. RELA^-/-^ HEK293T cells were transfected with a NF-κB Firefly luciferase reporter and thymidine-kinase Renilla luciferase reporter plasmid together with RELB and NFKB2 coding plasmids. Additional transfection of either an empty vector plasmid or containing the coding sequence of wt IKKα or mutants IKKα allowed the assessment of their own activity toward the activation of the non-canonical NF-κB pathway. **B**. Relative luciferase activity resulting from the transfection of an increasing dose of a plasmid coding IKKα WT, p.H142R, p.P190L, p.Y580C or p.Q422* (from 20 to 70 ng as indicated), co-transfected with RELB and p100. The NF-κB Firefly signal was normalized to the Renilla signal and then to transfection with RELB p100 only. An empty vector (EV) was co-transfected in each condition to reach a constant amount of plasmid. **C**. Western-blot of phosphorylated IKKα/β before and after immunoprecipitation of HA-tag IKKα transfected in HEK293T cells **D**. Western-blot of HA-tag IKKα after transfection of HEK293T cells for kinase assay. EV stands for empty-vector and WT for the transfection of IKKα WT. **E**. IKKα kinase activity after pulldown of the different alleles expressed as the relative luminescence of the wt allele or the variants to the empty vector. Each point represent one experiment.

To confirm the loss of kinase activity of the patient’s alleles, we transfected the variants in HEK293T cells and performed an immunoprecipitation using an anti-HA tag antibody (**Figure 3C-D**). We observed a complete loss of autophosphorylation of IKKα upon transfection as compared to the wt or the p.Y580C published variant (**Figure 3C**). A kinase activity was measured using an ADP-Glo Luciferase system (see methods). While we observed an increased kinase activity after pulling-down the wt IKKα, the activity was profoundly impaired for both studied variants while IKKα p.Y580C showed a normal activity (**Figure 3D-E**). This was also true by looking at the phosphorylation at serine 32 of a GST-tagged IκBα (**Figure 3F**). Finally, co-transfection of NIK and the different variants showed that, in contrast to wt IKKα, the variants were less able to induce the degradation of NIK, a phenomenon known to be linked to IKKα’s kinase activity which phosphorylates NIK to mediate its degradation in a retrocontrol loop (**Supplementary Figure 4A**) *(Razani et al., 2010)*. These experiments strongly supported that both the IKKα p.H142R and IKKα p.P190L proteins displayed a profound defect in kinase activity.

### Impaired non-canonical NF-κB pathway activation in patient’s cells

IKKα is involved in the phosphorylation of p100 that induces its processing into p52, leading to the translocation of a p52:RelB dimer in the nucleus *(Senftleben et al., 2001)*. LTβR (stimulated with LTα1β2), Fn14 (stimulated with Tweak) are receptor known to engage the non-canonical pathway*(Dejardin et al., 2002; Roos et al., 2010; Saitoh et al., 2003)*. We previously showed that both variants were kinase loss-of-function (**Figure 3, Supplementary Figure 4A**). In line with this, the cleavage of p100 to p52 was nearly abolished at different time points post-stimulation in patient’s SV40 fibroblasts, indicating an impaired activation of the non-canonical NF-κB pathway (**Figure 4A**). We showed that this observation was not due to an impaired interaction with NIK as assessed by co-immunoprecipitation after transfection (**Supplementary Figure 4B**). The same impairment in p100 cleavage was observed in anti-CD3 activated T cells from the patient (**Figure 4B**). This was associated to an impaired proliferation of activated T cells upon anti-CD3 stimulation, and an impaired activation assessed by the upregulation of CD69 (**Supplementary Figure 4A**). Accordingly, the translocation of RELB into the nucleus was nearly abolished in SV40-fibroblasts as assessed by western blots after LTα1β2, Tweak or TNFα stimulation or by EMSA experiments (**Figure 4C, Supplementary Figure 4B-C**). We also observed that *VCAM1* expression, a known target gene of the NF-κB dimer RELB:p52 in fibroblasts, was lower at the transcriptomic level upon LTα1β2 stimulation of SV40-fibroblast (**Supplementary Figure 4D**). These results were consistent with those reported with the homozygous p.Y580C variant *(Bainter et al., 2021)*. Altogether, we showed that the defective kinase activity of both the IKKα p.H142R and the IKKα p.P190L mutants resulted in a profound impairment of the non-canonical NF-κB pathway activation.

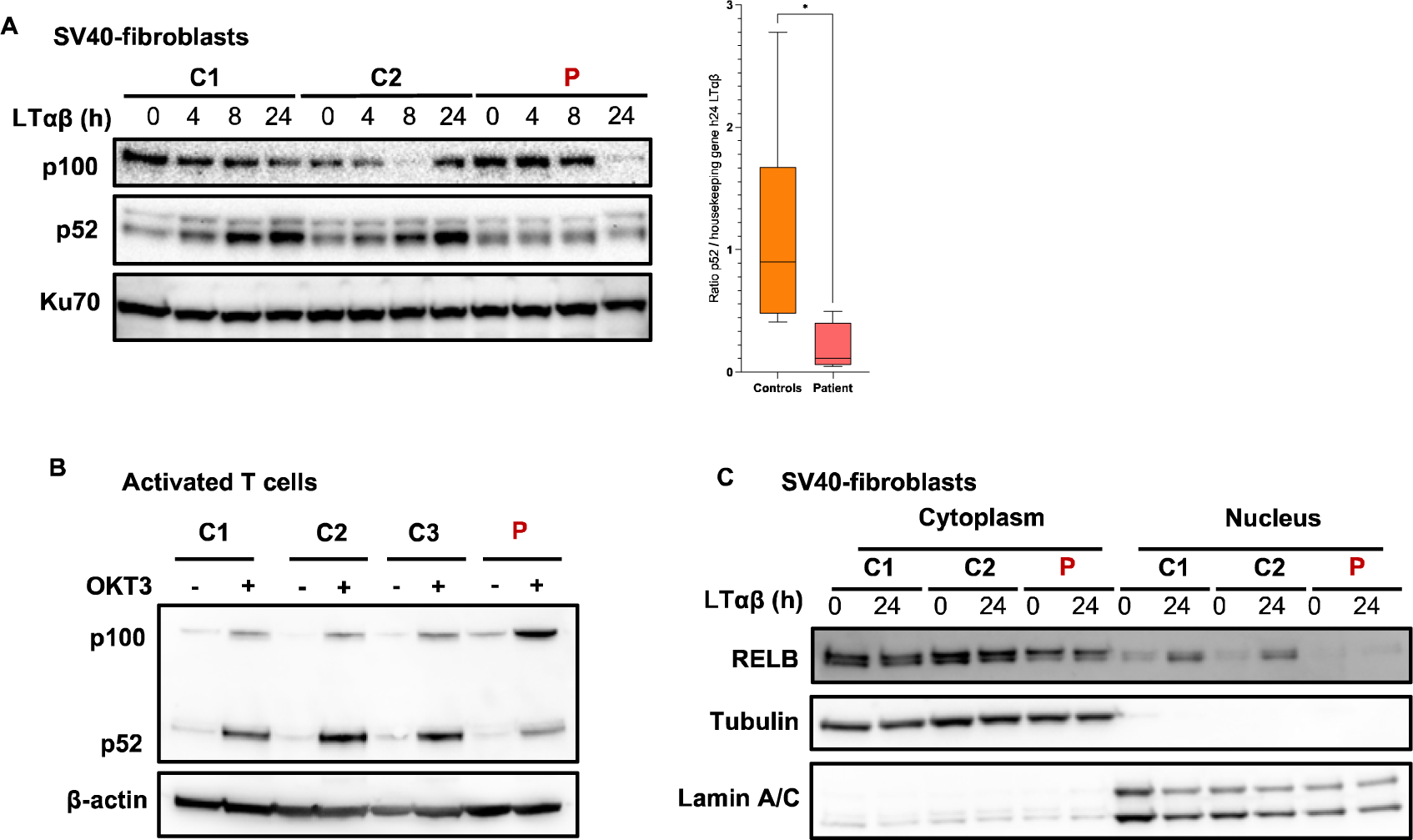
Figure 4.

**Figure 5.**
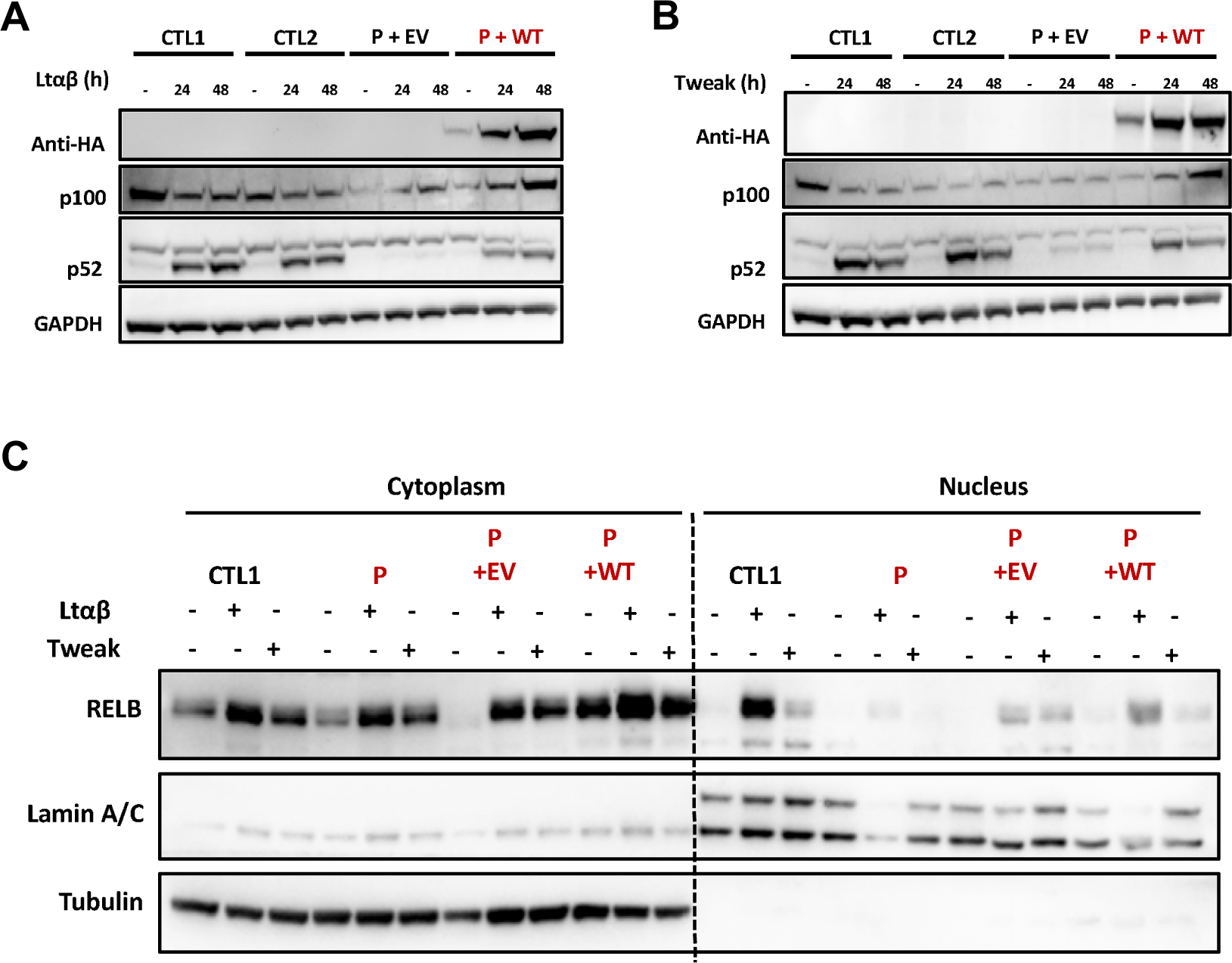
Reconstitution of patient cells with IKKα restored the non-canonical NF-κB defect *in vitro*. **A & B**. Patient cells were transduced with an EV (P+EV) lentivirus or containing the coding sequence of wt IKKα (P+WT). GFP positive cells were sorted and cell lysate after activation of the non-canonical pathway by LTαβ (**A**) or Tweak (**B**) were subjected to western blot. p100 and p52 were revealed using specific antibodies and GAPDH was used as a loading control. **C**. Patient cells were transduced with an EV (P+EV) lentivirus or containing the coding sequence of wt IKKα (P+WT). GFP positive cells were sorted. After activation with either LTαβ or Tweak, cytoplasmic and nuclear protein separation was performed and subjected to western blot. RELB was assessed using a specific C-terminal antibody to assess its nuclear translocation after activation in transduced patient’s cells compared to controls. Tubulin was used as a loading control of cytoplasmic extract while Lamin A/C was used for nuclear extracts.

### Reconstitution of patient’s SV40-fibroblasts restores a normal non-canonical NF-κB phenotype

To further confirm that the identified *CHUK* variants were responsible for the observed *in vitro* phenotype, we transduced fibroblasts from the patient with an empty lentivirus or a lentivirus containing the wt *CHUK* coding sequence. The transduced cells were stimulated by LTα1β2 or Tweak and we assessed the cleavage of p100. We still observed the impaired cleavage of p100 to p52 as well as the impaired RELB nuclear translocation after 24 or 48h of LTα1β2 or Tweak stimulation in the cells transduced with the empty lentivirus (**Figure 5**). In contrast, transduction of the wt IKKα rescued the p100 cleavage as well as the nuclear translocation of RELB as compared to healthy control SV40-fibroblasts (**Figure 5A-C**). These results supported the causality of the two *CHUK* variants and confirmed an autosomal recessive form of IKKα deficiency.

### Partially impaired transcription of canonical NF-κB target genes

IKKα belongs to the NEMO complex along with the catalytic subunit IKKβ and the regulatory subunit IKKγ. Although previous publications showed that IKKα was dispensable in the IKK complex to phosphorylate IκBα *(Hu et al., 2001, 1999; Polley et al., 2016)*, a major checkpoint of the canonical NF-κB pathway activation, we had several arguments in favor of a partial impairment of the canonical pathway activation in cells from the patient. First, the clinical phenotype of the patient, suffering from ectodermal dysplasia, and the results of the immunophenotype, showing an excess of naïve CD4+ and CD8+ T cells, are clinical arguments for the involvement of the canonical NF-κB pathway *(Puel et al., 2004)*. In addition, contrary to what was found by Bainter et al. with the *CHUK* p.Y580C homozygous variant, affecting only the non-canonical pathway, induction of *VCAM1* as assessed by RTqPCR was low in patient’s SV40-fibroblasts stimulated with TNFα *(Bainter et al., 2021)* (**Supplementary Figure 5A**). Lastly, TNFα stimulation of the patient’s SV40 fibroblasts led to a lower induction of RELB protein expression, a known target gene of the canonical NF-κB pathway (**Supplementary Figure 4A**). We first assessed the phosphorylation of p65 and IκBα along with IκBα degradation that were not affected in patient’s cells, suggesting a normal activity of the NEMO complex and confirming in human the redundant role of the kinase activity of IKKα in this complex (**Figure 6A**). Although we did not see any difference in the cytosolic phosphorylation cascade, we observed that the nuclear translocation of p65 was diminished upon TNFα stimulation (**Figure 5B**). We thus assumed that the IκΒδ inhibitory activity of p100 could be at the origin of this mild translocation defect of p65 given that the patient’s cells are greatly defective for the cleavage of p100 *(Tao et al., 2014; Mitchell et al., 2023)*. We compared the translocation of p65 into the nucleus of SV40-fibroblasts from the patient with that from a previously reported patient carrying the heterozygous *NFKB2* variant p.R853* *(Le Voyer et al., 2023)*, resulting in the loss of p100 cleavage in p52 and accumulation of p100. There was still a mild reduction in p65 translocation for the *CHUK* patient while it was not observed in the case of the *NFKB2* patient (**Supplementary Figure 5B**).

**Figure 6.**
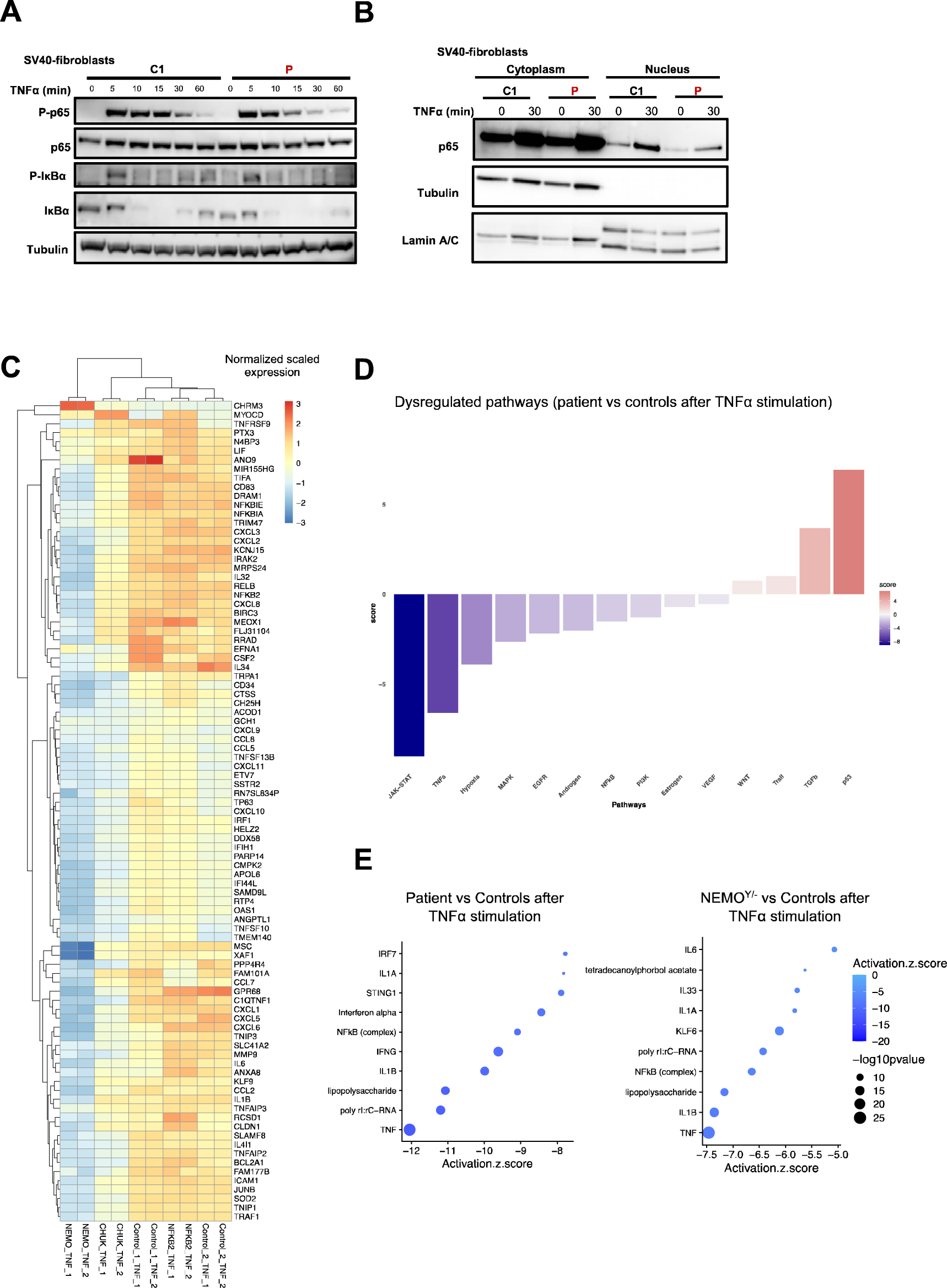
Exploration of the canonical NF-κB signaling in patient cells. **A**. Western-blot assessing the quantity of phospho-p65 (S536), p65, phospho-IκBα (S32) and IκBα on cell lysates from SV40-fibroblasts stimulated with TNFα at indicated time points. Tubulin was used as a loading control. **B**. Western-blot of cytoplasmic and nuclear protein extracts from SV40 fibroblasts activated with TNFα for 30 minutes assessing the translocation of p65/RELA in the nucleus. Tubulin was used as a loading control of cytoplasmic extract while Lamin A/C was used for nuclear extracts. **C.** Heatmap showing the normalized and scaled expression (vst) of genes significantly upregulated in control SV40-fibroblasts after stimulation with TNFα 10 ng/mL for 6 hours. Each column represents a patient or a control. RNA sequencing has been performed in biological duplicates for each control or patient (as indicated by _1 or _2 at the end of each column name). **D.** Pathway enrichment score of differentially expressed genes between the patient and controls SV40-fibroblasts stimulated with TNFα. A negative blue bar indicates a downregulation of the indicated pathway based on the PROGENy database while a red bar indicates an upregulated pathway. **E**. Z-score of the different “molecules” predicted to be inhibited (top 10 based on the z-score) based on Ingenuity Pathway Analysis (IPA, QIAGEN Inc) for the mentioned conditions. A negative score indicate that the molecule is predicted to be inhibited based on the differentially expressed genes that have been processed.

To go deeper in the characterization of the canonical NF-κB pathway, we performed a bulk RNA sequencing of SV40-fibroblasts from the patient and controls before and after 6 hours of stimulation with TNFα or Poly I:C (TLR3 agonist) two potent activators of the canonical NF-κB pathway (**Figure 6C-E**). We also included *NEMO*^Y/-^ SV40-fibroblasts (*NEMO* null), known to be unresponsive to TNFα stimulation *(Smahi et al., 2000)*, as well as the SV40-fibroblasts from a patient carrying the heterozygous *NFKB2* p.R853* variant *(Le Voyer et al., 2023)*. Upon TNFα stimulation, TNFα induced-genes or genes from the TNFα signaling via NF-κB signature (MSIgDB, **supplementary Figure 5C**) were significantly less upregulated in fibroblasts from the patient than in fibroblasts from controls (**Figure 6C**). Accordingly, pathway activity inference based on previous knowledge (PROGENy) showed a profound defect in the activation of the TNFα signaling (**Figure 6D**) *(Badia-I-Mompel et al., 2022)*. This was confirmed by Ingenuity Pathway Analysis (QIAGEN) showing that TNFα, IL1β and lipopolysaccharide (LPS), 3 potent activators of the canonical NF-κB pathway, were in the top 3 molecules predicted to be inhibited based on the differentially expressed genes (DEGs) between the patient’ cells and control cells (**Figure 6E**). Although DEGs analysis for the NF-κB target genes induced after TLR3 stimulation suggested similar activation between the patient and controls (**Supplementary Figure 6A**), a list of genes was consistently downregulated after both TNFα and TLR3 stimulation (*RELB*, *NFKB2*, *NFKBIA*, *ICAM1*, *BIRC3* among others, **Supplementary Figure 6B**). Moreover, molecules predicted to be inhibited were mostly NF-κB inducers (**Supplementary Figure 6C**). Of note, IRF3 target genes after TLR3 stimulation were normally upregulated in the patient cells (**Supplementary Figure 6D**). In contrast to the IKKa patient’ cells, the NFKB2 p.R853* patient’s cells were in the range of controls both after TNFα and TLR3 stimulation for NF-κB target genes (**Figure 6C**) while NEMO null cells were unable or less able to upregulate genes upon TNFα and TLR3 stimulation, as previously reported (**Figure 6C, Supplementary Figure 5**)*(Audry et al., 2011)*.

Altogether, these results suggested that the two missense variants in IΚΚα negatively impacted the canonical NF-κB pathway, although to a lesser extent than a complete loss-of-activation as in NEMO *null* cells, without affecting the cytosolic function of the NEMO complex.

### Neutralizing autoantibodies against type I interferons are detectable in the patient serum

This patient suffered from a severe deficiency of the non-canonical NF-κB pathway. Recently, neutralizing autoantibodies against type I interferon were described as a hallmark of non-canonical NF-κB deficiency in patients with p52 LOF/ p100 IκBδ GOF *NFKB2* variants, bi-allelic loss-of-function *NIK* and *RELB* variants *(Le Voyer et al., 2023; Bodansky et al., 2023)*. Using a previously described luciferase assay, we questioned the presence of such autoantibodies in autosomal recessive IKKα deficiency *(Le Voyer et al., 2023; Bastard et al., 2021)*. Interestingly, serum from the patient (5 years before her death, well before the SARS-CoV-2 pandemic) neutralized IFN-α2 both at 10 ng/mL (**Figure 7A**) and 100 pg/mL (more physiological concentration, **Figure 7B**) and IFN-ω at 100 pg/mL but did not neutralize IFN-β (**Figure 7A-B**).

**Figure 7.**
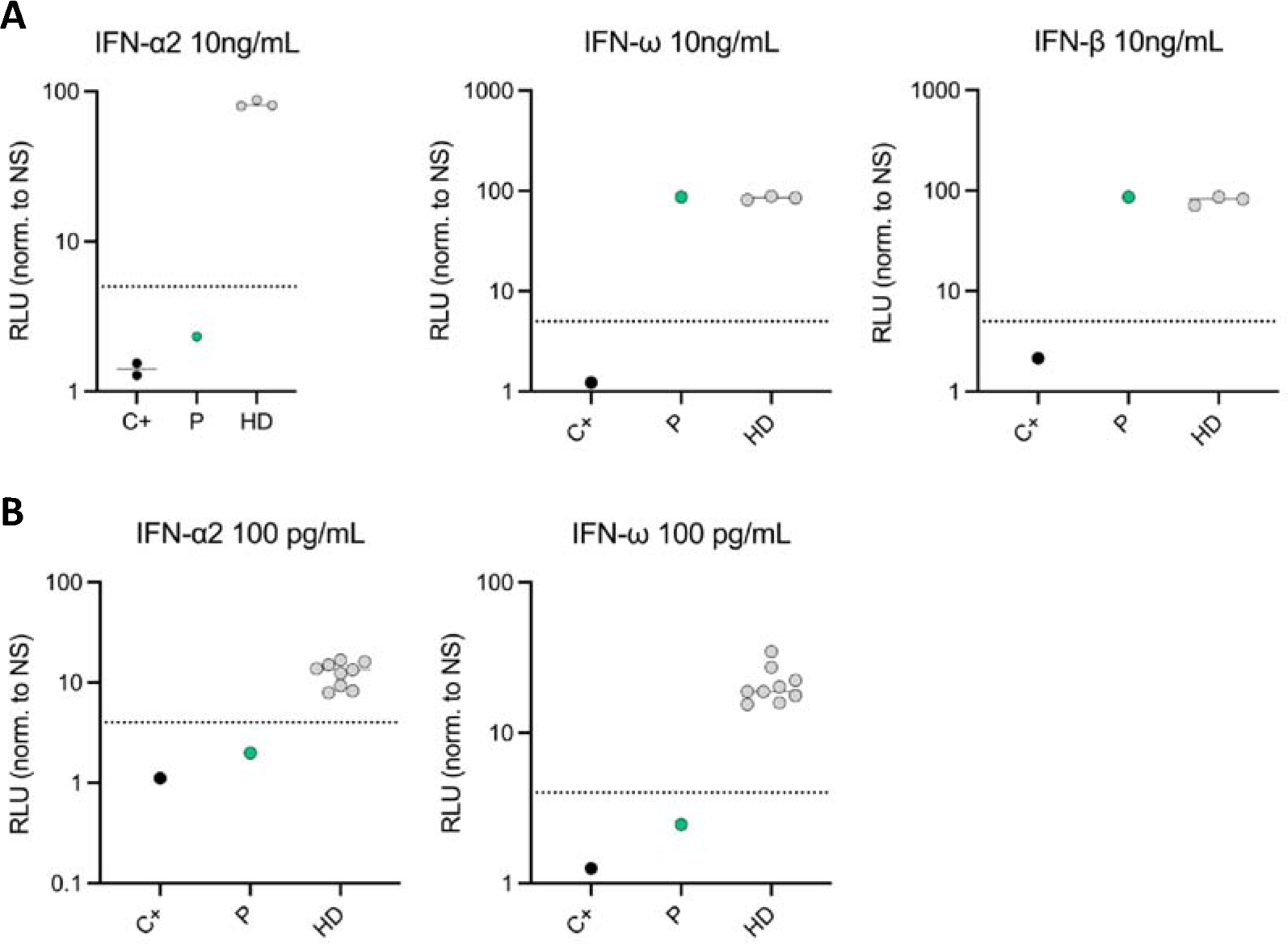
Neutralizing autoantibodies against type I IFNs. **A**. Luciferase-based neutralization assay to detect auto-Abs neutralizing 10 ng/mL IFNα2, IFN-ω or IFN-β, in an individual previously known to have neutralizing anti-IFN-I auto-Abs (C+), healthy donors (HD), and the patient and the patient with the variants in *CHUK* (P). **B**. Luciferase-based neutralization assay to detect auto-Abs neutralizing 100 pg/mL IFNα2, IFN-ω in an individual previously known to have neutralizing anti-IFN-I auto-Abs (C+), healthy donors (HD), and the patient with the variants in *CHUK* (P). Relative luciferase unit (RLU) is calculated by normalizing Firefly luciferase activity against the Renilla luciferase activity, before normalization against non-stimulated conditions. An induction factor of less than five relative to non-stimulated conditions was considered to correspond to neutralizing activity (dashed line).

## Discussion

In this study, we report the first compound heterozygous loss-of-function mutation in *CHUK* leading to a severe loss of IKKα kinase activity. In humans, the complete absence of the protein IKKα (*CHUK null*) has been shown to be lethal *in uter*o, a phenotype reminiscent of complete IKKα deficiency (IKKα^-/-^) in mice which die early after birth *(Hu et al., 2001; Takeda et al., 1999; Lahtela et al., 2010; Hu et al., 1999)*. Only one patient presenting with a combined immunodeficiency and a homozygous missense mutation in *CHUK* was described up to now *(Bainter et al., 2021)*. This mutation (p.Y580C) changed a residue in the helix-loop-helix domain of IKKα and was shown to abrogate the interaction between IKKα and its activating kinase NIK. Consequently, cells from that patient exhibited a profound defect of the non-canonical NF-κB pathway activation with an impaired cleavage of p100 to p52 despite a normal kinase activity of the mutant *in vitro*. The phenotype was reproduced in a mouse model. Importantly, the canonical NF-κB pathway was not reported to be affected in the patient carrying the homozygous IKKα p.Y580C *(Bainter et al., 2021)*.

The patient herein studied suffered from a syndromic combined immunodeficiency including hypogammaglobulinemia, recurrent infections, digestive and liver inflammation, along with bone malformation and ectodermal dysplasia. Two heterozygous private missense mutations modifying highly conserved residues in the kinase domain of IKKα were identified. We showed that both alleles were kinase loss-of-function *in vitro* and that this was leading to a complete deficiency in the activation of the non-canonical NF-κB pathway in the patients’ cells. These results were consistent with the patient’s phenotype which included hypogammaglobulinemia, low B-cells, absence of memory B cells and plasmablasts, and related recurrent infections as well as the presence of autoantibodies against type I IFNs. All these manifestations were previously reported in other defects affecting the non-canonical NF-κB pathway *(Willmann et al., 2014; Schlechter et al., 2017; Sharfe et al., 2015; Le Voyer et al., 2023; Bainter et al., 2021)*. Moreover, mice knocked-in for a kinase dead IKKα (IKKα p.K44A, IKKα^KA/KA^) showed an early arrest of B cell development associated with an impaired non-canonical NF-κB signaling and impaired formation of secondary lymphoid organs *(Balkhi et al., 2012)*. We therefore concluded that this patient suffered from a very severe deficiency of IKKα kinase activity.

Interestingly, we also observed a partial defect in the transcription of canonical target genes in the SV40-fibroblasts from the patient upon TNFα stimulation. According to previous publications showing a redundant role of IKKα in the NEMO complex, we did not observe differences in the phosphorylation or degradation of IκBα in cells from the patient *(Hu et al., 1999)*. In contrast, bulk RNA sequencing of SV40-fibroblasts before and after stimulation with TNFα or poly I:C showed a partial defect in the transcription of canonical NF-κB target genes. Whether this result was linked to an excess of the uncleaved p100 exerting an IκΒδ inhibitory effect on p65-containing dimers was a possibility *(Tao et al., 2014; Razani et al., 2010)*. In the mice model IKKα^KA/KA^ it was already described that p65 was less present in the nucleus in bone marrow B cells and the authors suggested that p100 could inhibit its translocation *(Balkhi et al., 2012)*. Consistently, we observed a mild defect in the nuclear translocation of p65 after TNFα stimulation. In patients with autosomal recessive NIK deficiency, the canonical pathway was impacted but the experiments did not specifically look at the role of the p100 excess *(Willmann et al., 2014)*. In *NFKB2* deficiency, the relative excess of p100 is a hallmark of the mutations affecting the C-terminal processing inhibitory domain (PID)*(Le Voyer et al., 2023)*. In an ectopic expression model of those *NFkB2* mutations, it was shown that p100 accumulation was associated to a cytoplasmic retention of p65 *(Lee et al., 2014)*. In our report, cells from an *NFKB2* mutated patient (p52 loss-of-function/p100 gain-of-function) did not exhibit any defect of the canonical NF-κB pathway activation as assessed by bulk RNA sequencing after TNFα or poly I:C stimulation. Moreover, we observed a normal p65 nuclear translocation upon TNFα stimulation. Therefore, the defect observed for particular canonical NF-κB target genes activation observed in the patient carrying two IKKα LOF mutations could be a direct consequence of the defective kinase activity rather than the relative excess of p100 or an association of both mechanisms. A possible explanation for this observation may rely in the nuclear role of IKKα *(Swaminathan, 2003)*. It was previously shown that IKKα shuttle into the nucleus where it phosphorylates histone H3 on Serine 10 thereby promoting the transcription of NF-κB target genes *(Anest et al., 2003, 2004; Yamamoto et al., 2003)*. The defective kinase activity observed for both mutants could therefore lead to less histone H3 phosphorylation upon TNFα stimulation, and potentially dampening the transcription of some NF-κB target genes as observed in our RNAseq experiment. Whether this also explained the reduction in the activation of the JAK-STAT pathway as observed by inference analysis or if this was an undirect consequence of the lower level of transcription of IL-6 and IFNγ remains to be determined. In any case, the defective activation of the NF-κB canonical pathway could account for the phenotype of the patient who displayed many features of canonical defect such as dental enamel abnormalities, a skin disease resembling ectodermal dysplasia and elevated naive CD4+ and CD8+ T cell, clinical and biological features known to occur in patient with autosomal dominant gain-of-function mutations in *NFKBIA* or hypomorphic X-linked mutations in *NEMO (Smahi et al., 2000; Courtois et al., 2003; Courtois and Smahi)*. Of note, *TP63,* the gene known to be mutated in Hay-Wells syndrome for which the patient was first sequenced due to similar clinical skin and bones phenotype was found downregulated in patient fibroblasts upon TNFα stimulation in our RNAseq experiment. Whether this result could explain the severity of the skin and bone phenotype was beyond the scope of this study and would require further investigations.

Intriguingly, the patient died from complications related to a diffuse large B-cell lymphoma (DLBCL) of the lung, a malignancy frequently driven by the excess of NF-κB activation *(Eluard et al., 2022)*. It was thus surprising that this type of lymphoma developed in the context of this IKKα deficiency. However, a similar, although rare, observation was made in other inborn errors of immunity such as monoallelic *NFKB2* deficiency, with only one case of non-Hodgkin lymphoma (non-EBV related) *(Klemann et al., 2019)*. Interestingly, mice models have suggested a role of IKKα in preventing skin tumor development by the suppression of EGFR and c-Myc pathways while another model showed that mice IKKα^KA/KA^ were prone to develop systemic inflammation and lung tumors (lung squamous cell carcinomas) *(Descargues et al., 2008; Xiao et al., 2013)*. Whether similar mechanisms (e-g non-NF-κB roles of IKKα) explain the development of a B-cell lymphoma in this patient remain to be determined and the contribution of additional somatic genetic drivers has not been investigated *(Leveille and Johnson, 2021)*. We also could not exclude the contribution of a defective immunosurveillance in this disease as the patient’s T cells failed to proliferate *in vitro* upon non-specific TCR stimulation *(Schreiber et al., 2011; Afshar-Sterle et al., 2014)*. In contrast, the patient did not suffer from gross cellular immunity deficiency except for the onychomycosis that could be linked to a defect in Th17 as previously described in the other patient with IKKα deficiency *(Bainter et al., 2021)*. Interestingly, the patient herein described and the other patient with the homozygous p.Y580C variant both had liver complications and villous atrophy, the latter one being more severe as it required liver transplantation *(Bainter et al., 2021)*. Such clinical manifestations were already described in NIK and RELB deficiencies *(Willmann et al., 2014; Ovadia et al., 2017)*. A possibility could be that the non-canonical NF-κB activation defect was at the origin of a global immune dysregulation and infiltration of organs with autoreactive T cells although this was not demonstrated. In line with this was the autoimmunity and uncharacterized immune dysregulations with lymphocytic organ infiltration widely reported in *NFKB2* mutated patients *(Klemann et al., 2019)*. Among several possible mechanisms, the break of central tolerance due to the impaired expression of Aire by medullary epithelial thymic cells (mTECs) as previously described in mice and recently in human was an appealing hypothesis *(Le Voyer et al., 2023; Zhu et al., 2006)*. Accordingly, the studied patient displayed neutralizing autoantibodies against type I IFNs as recently shown in human with other germline deleterious mutations of the non-canonical NF-κB pathway *(Le Voyer et al., 2023; Bodansky et al., 2023)*. Another possibility could be that the patient herein studied suffered from a recently described entity called EVAH for enteric viral-associated hepatitis where chronic infections with enteric viruses in immunodeficient patients were associated to chronic hepatitis *(Riller et al., 2023; Fourgeaud et al., 2023; Bucciol et al., 2018)*

Overall, we described the first human bi-allelic mutations in *CHUK* leading to a severe deficiency of the IKKα kinase activity. As expected, the NIK-IKKα dependent non-canonical NF-κB pathway was severely compromised. Our findings also showed that induction of particular canonical NF-κB target genes was partially impaired which may explain the severity of the patient’s phenotype, including its skin abnormalities, and emphasize a role of IKKα kinase activity in the human canonical NF-κB pathway activation. The study of this single patient emphasized that a severe deficiency of IKKα kinase activity was compatible with life in human, although associated to immunodeficiency and malformations, while the complete loss of protein expression was lethal *(Lahtela et al., 2010)*.

## Data Availability

All data produced in the present study are available upon reasonable request to the authors

## Acknowledgements

We would like to thank the core facilities of the Imagine Institute (bioinformatics, genomics, flow cytometry) and the Flow Cytometry Platform of the Pitié-Salpêtrière Hospital (CyPS) for CyTOF acquisition and technical advice. This work was supported by the Institut National de la Santé et de la Recherche Médicale (INSERM) and by government grants managed by the Agence National de la Recherche as part of the “Investment for the Future” program (Institut Hospitalo-Universitaire Imagine, grant no. ANR-10-IAHU-01; Recherche Hospitalo-Universitaire, grant no. ANR-18-RHUS-0010), the Centre de Référence Déficits Immunitaires Héréditaires (CEREDIH), the Agence National de la Recherche (grant nos. ANR-18-CE17-0001 “Action”; and ANR-22-CE15-0047-02 “BREAK-ITP”), the Fondation pour la recherche Médicale (FRM: EQU202103012670). The Laboratory of Human Genetics of Infectious Diseases is supported by the Howard Hughes Medical Institute, the Rockefeller University, the St Giles Foundation, the National Institutes of Health (NIH) (R01AI127564), the Fisher Center for Alzheimer’s Research Foundation, the Meyer Foundation, the JBP Foundation, the French National Research Agency (ANR), the Integrative Biology of Emerging Infectious Diseases Laboratory of Excellence (ANR-10-LABX-62-IBEID), ANR GENVIR (ANR-20-CE93-003), ANR AAILC (ANR-21-LIBA-0002) and ANR AI2D (ANR-22-CE15-0046) projects, the ANR-RHU programme (ANR-21-RHUS-08-COVIFERON), the HORIZON-HLTH-2021-DISEASE-04 programme under grant agreement 101057100 (UNDINE), the Square Foundation, Grandir—Fonds de solidarité pour l’enfance, the SCOR Corporate Foundation for Science, the Battersea & Bowery Advisory Group, Stavros Niarchos Foundation (SNF) as part of its grant to the SNF Institute for Global Infectious Disease Research at The Rockefeller University, William E. Ford, General Atlantic’s Chairman and Chief Executive Officer, Gabriel Caillaux, General Atlantic’s Co-President, Managing Director and Head of business in EMEA, and the General Atlantic Foundation, the French Ministry of Higher Education, Research, and Innovation (MESRI-COVID-19), Institut National de la Santé et de la Recherche Médicale (INSERM) and of Paris Cité University. Tom Le Voyer (TLV) was supported by the Bettencourt-Schueller Foundation and the MD-PhD program of Imagine Institute. This work was also supported by grants from Institut National du Cancer, Fondation ARC pour la Recherche sur le Cancer, University Paris Cité, 12 Rounds contre le Cancer (V.B.), and Doctoral funding from the French Ministry of Higher Education and Research (T.B.). I.M. is a senior clinical investigator at FWO Vlaanderen and is supported by the Jeffrey Modell Foundation and by FWO Grant G0B5120N. This work is supported by ERN-RITA. QR received an Institut Imagine MD-PhD fellowship and a Société Nationale Française de Médecine Interne (SNFMI) fellowship.

## Authorship contributions

Study design: F.R-L., B.N., Q.R., A.F., A.P.; Experiments: Q.R., B.S.,C.C., D.H-N., T.L.V.,O.P.,M-C.S., M.J., T.B., C.B., C.Br., V.M., M.R.R.,C.R., J-C.D, A.M., V.M., C.Ro., L.D.; Generation of bulk RNA sequencing dataset: Q.R., C.M., C.Bole., N.Ca.; Bioinformatic analysis of RNA-seq dataset and CyTOF: A.C., Q.R.; Writing original draft: Q.R., F.R-L.; Writing Review and editings: A.F., I.M., V.B., J-L.C., A.P., B.N., E.D.; Resources: C.Bo., B.N., L.B., I.M.; All authors reviewed and approved the final version.

## Material and methods

### Study participants

The study was approved by the “Comité d’éthique de la recherche Assistance Publique Hopitaux de Paris” (Institutional Review Bord Number #00011928, Paris, France). The study was performed in accordance with the 1975 Declaration of Helsinki and subsequent revisions. The biological samples are part of Inserm and Imagine institute collection declared to the French Ministère de la recherche (Access to the Human Body Element Conservation Management Application no. DC-2020-3994, Paris, France).

### Genetic analysis

Next-generation sequencing was previously described in the publication of one mutation of the patient (36). Sanger sequencing was performed to confirm the next-generation sequencing results and to analyze complementary DNA and genomic DNA from primary cell lines of the patient. Purified PCR products were directly sequenced using BigDye Terminators (version 1.1) and a 3500xL Genetic Analyzer (Applied Biosystems).

### Primary fibroblasts isolation

Skin biopsy was separated in two parts by using scalpel and introduce in a 25 cm^2^ culture flask (Fisher Scientific). After 15 min without medium at 37°C in a humidified 5% CO2 incubator, skin biopsies attached to the plastic of the flask and culture medium was added (RPMI 1640 from Life Technologies supplemented with 20% Fetal Bovin Serum (FBS, Life Technologies), 1% penicillin-streptomycin (PS, Thermofischer), 1% amphotericin B (Thermofischer), 50 ug/mL gentamicin (Thermofischer). Fibroblasts usually starts to grow after 1 or 2 weeks of culture and are trypsinized (Trypsin, Thermofischer) and cultured in 25 cm^2^ culture flasks (passage 1) and then in 75 cm^2^ culture flasks (passage 2). After these two passages, cells are frozen in FBS 10% dimethylsulfoxide (DMSO, Thermofischer).

### Cell culture

SV40-fibroblasts, HEK293T and RELA^-/-^ HEK293T cells were maintained in DMEM (Life Technologies) supplemented with 10% fetal bovine serum (FBS, Thermo Scientific Gibco) and 1% penicillin/streptomycin (PS, Thermo Scientific). B-EBV cell lines were maintained in complete RPMI 1640 (Life Technologies RPMI supplemented with 10% FBS, 1% PS). T cells activation was carried out from the PBMC of patients or of healthy donors. T cells were activated by binding CD3/CD28 receptors with Dynabeads (Thermofisher, #11141D). Cells were cultured at 1.10^6^/mL/10 μL of Dynabeads in complete Panserin medium (Panserin 401 medium (PAN BIOTECH, # P04-710401), supplemented with 5% human AB serum, 1% pennicilin/streptomycin (Thermofisher, 15140122) and 1% L-glutamine (Thermofisher, 25030081) and filtered with a unit Nalgene Rapid-Flow single-use sterile filtration unit with PES membrane (Thermofisher, # 568-0010). After 3 days of culture, cells were centrifuged on Ficoll and then recultured in complete Panserin with interleukin-2 (IL-2) at 100 international units/mL (U/mL). Every two days, cell expansion was continued by adding Panserin complete with IL-2.

### Transformation of primary fibroblasts with SV40

Primary fibroblasts were immortalized by lipofection of 2.5 ug of pBSSVD2005 (Addgene #21826) coding the large-T antigen of Simian virus-40, using Lipofectamine 2000 as previously described in the methods. After 3 to 4 cycles of cultures, fibroblasts changed their shape and growing rate and were considered transformed and called SV40-fibroblasts.

### CRISPRCas9 in HEK293T cells

CRISPR-Cas9 technology was used to knock-out RELA from WT HEK293T cells. RNA guides were designed using the online tool CRISPOR. Single guides cDNA were cloned into MLM3636 plasmid (Addgene plasmid #43860). Sequences of the plasmids were verified by Sanger sequencing. WT HEK293T cells were transfected using lipofectamine 2000 (Life Technologies) with the plasmid MLM3636 containing the cDNA sequence of the single-guide and a plasmid coding the Cas9 (pCas9_GFP, Addgene plasmid #44719). Forty-eight hours later, GFP-positive cells were single-cell sorted in flat-bottom 96 wells plates (Falcon). Growing clones of transfected HEK293T cells were screened by Sanger sequencing for genomic alteration around the single-guide target. Homozygous clones for frameshift deletion or insertion in *RELA* were selected and subjected to western-blot for the N-terminal part of p65. Loss-of-expression clones were selected for further experiments. Response to TNFα was null in NF-κB luciferase experiments. These cells are called RELA KO HEK293T cells in the manuscript. DNA sequence of the single guides are as follow: Fw 5’ ACA CCG CTT TCC TAC AAG CTC GTG GG 3’; Rv 5’ AAA ACC CAC GAG CTT GTA GGA AAG CG 3’.

### Subcloning in mammalian expression vectors

*CHUK* complementary DNA (cDNA) was amplified using primers encompassing the 5’UTR (forward) and the 3’UTR (Reverse) from control PBMC cDNA. The agarose gel band at the expected size was purified and amplified by using primers adding a restriction site at each end of the coding sequence (from ATG to TAA). The purified band was subcloned into the mammalian expression vector pCMV-HA-tagged (Clontech). Directed mutagenesis was performed following the Q5-site directed mutagenesis kit protocol (NEB).

pCMV-MYK-tagged NIK and pEF1A-p100 were obtained by subcloning NIK and p100 from available backbones (pCMV4-p100 Addgene #23287, EGFP-NIK Addgene #111197, respectively) using the NEBuilder HiFi DNA Assembly Cloning kit following manufacturer instructions (NEB). pCMV6-RELB was kindly provided by the team of Pr. Jean-Laurent Casanova. Sanger sequencing was used to verify the sequence of the plasmids from the promoter to the end of the insert after subcloning and directed mutagenesis.

HA-CHUK coding sequence was also transferred to a lentiviral plasmid backbone (pMSCV-MCS-EF1-GFP-T2A-Puro) using the NEBuilder HiFi DNA Assembly Cloning kit. All plasmid production were performed by transforming DH5α E.Coli competent cells (NEB) or Stbl3 for the lentivirus plasmids (NEB). Extraction and purification of plasmid DNA was performed on PureLink HiPure Plasmid Midiprep Kit (ThermoFischer) or Wizard Plus SV Minipreps DNA Purification Systems (Promega).

### Transfection of HEK293T cells

HEK293T cells were seeded in plates 24h before the transfection. WT or RELA^-/-^ HEK293T cells were transiently transfected using Lipofectamine 2000 (Life Technologies) with the different plasmids as indicated in the figures (*CHUK*, *RELB*, *p100*, *NIK*). The DNA and Lipofectamine 2000 dilution were performed in optiMEM (Life Technologies). Twenty-four or forty-eight hours later, cells were harvested to perform western-blot or luciferase assays.

### Non-canonical NF-κB pathway luciferase in HEK293T cells

To activate specifically the non-canonical pathway in an ectopic expression model of *CHUK* we used the RELA^-/-^ HEK293T cells. RELA^-/-^ HEK293T cells were seeded in 96 flat-bottom compatible with luciferase reading plates (Greiner 655098) and transfected twenty-four hours later by using Lipofectamine 2000. Cells were transfected with pCMV-HA-tagged EV or *CHUK* WT or mutants, pEF1A-P100 WT and pCMV6-RELB depending on the experiments and two reporter plasmids, one Firefly Luciferase reporting NF-κΒ activity pGL4.32[luc2P/NF-κB-RE/Hygro] (Promega) and one pTK Renilla Luciferase as an internal control (ThermoFischer Scientific). Forty-eight hours later, luciferase signal was revealed and measured with the Dual-Glo Luciferase Assay System (Promega). The quantity of plasmid was kept constant in all conditions by adding the remaining dose of empty vector.

### Co-immunoprecipitation between NIK and IKKα

For the co-immunoprecipitation NIK/IKKα experiments, HEK 293T cells were transfected with a Flag-NIK kinase dead expression vector along with a vector encoding HA-IKKα either WT, H142R, P190L. Two days post-transfection, the cells were lysed (50 mM Tris pH 7,5, 1% NP-40, 1 mM EDTA, 6 mM EGTA, 120 mM NaCl, 15 mM sodium pyrophosphate, PhosSTOP (Sigma 4906845001) and Compete (Sigma 4906837001) and 20 mM NEM (N-ethylmaleimide, Sigma E1271)) and the protein extract was subjected to an immunoprecipitation with either control IgG-beads (Sigma A0919) or Anti-Flag-M2 beads (Sigma F2426). Half of the immunoprecipitates were used for an immunoblot for detecting Flag-NIK and the other half for the detection of HA-IKKα.

### Kinase assay

HEK293T cells were transfected with pCMV-HA EV, pCMV-HA CHUK p.H142R, p.P190L or p.Y580C. After 24h, cells were lysed in cold lysis buffer (containing Tris HCl pH 7 50mM, NaCl 150 mM, EDTA 1 mM, Triton 100-X 1%) and proteins were subjected to immunoprecipitation using an anti-HA tag monoclonal antibody (Sepharose Bead Conjugate, Cell Signaling clone C29F4) following manufacturer instructions, overnight at 4°C. The following day, sepharose beads were washed five times in cold lysis buffer and two times in room temperature 1X kinase buffer (Promega IKKα Kinase Enzyme System, V4068). Kinase reaction was performed for 1h at room temperature by adding 2X kinase reaction buffer containing ATP and IKKtide as a substrate on the sepharose pellet. The supernatant was used to measure the kinase activity using the ADP-Glo Luciferase-based kinase assay following manufacturer instructions (Promega V6930). Sepharose beads were washed again two times in lysis buffer and subjected to immunoblot after denaturation. Results are normalized to the empty vector condition.

### Kinase assay using GST-IκBα

For the kinase assay of IKKα, HEK 293T cells were transfected with a vector encoding HA-IKKα (WT, H142R, P190L or Y580C) or an Empty Vector (EV). Two days post-transfection, the cells were lysed (50 mM Tris pH 7,5, 1% NP-40, 1 mM EDTA, 6 mM EGTA, 120 mM NaCl, 15 mM sodium pyrophosphate, PhosSTOP (Sigma 4906845001) and Compete (Sigma 4906837001) and the protein extract was subjected to an immunoprecipitation with an anti-HA AB (BioLegend 901514) coupled to A/G Agarose beads (Santa-Cruz SC-2003). Kinase reaction was performed for 1h at 30°C in presence of 200ng GST-IκBα in a kinase assay buffer (20mM HEPES pH 7,5, 10mM MgCl2, 2mM DTT, 100uM ATP, PhosSTOP (Sigma 4906845001) and Compete (Sigma 4906837001). Immunoblots were then performed for the detection of p-IκBα S32

### Lentiviral production

HEK293T Lenti-X cells were cultured in DMEM 10% FBS 1% PS. The day before the experiment, cells were seeded in 75 cm^2^ flasks (Falcon) for cell culture. At D0, cells were transfected by Lipofectamine 2000 with 10 ug of the plasmid containing the coding sequence of HA-CHUK (with mutation or not, pMSCV-HA-CHUK-EF1A-GFP-T2A-Puro), 7.5 ug of psPAX2 (Addgene plasmid #12260) and 5 ug of pMD2.G (Addgene plasmid #12259). Sixty hours later, the media was collected, centrifuge at 3000 rpm at 4°C for 10 minutes to pellet cell debris. The supernatant was filtered through a 0.22 um low protein binding membrane (Millipore) and aliquots were stored at –80°C before use.

### Lentiviral transduction

SV40-fibroblasts were seeded in 6 well plates (Falcon) 24h before the transduction to reach 70 to 90% confluence. The day of transduction 1 mL of lentiviral production was added to 500 uL of optiMEM (Life Technologies) and transferred in each well of fibroblasts (either SV40-fibroblasts from the patient or healthy control SV40-fibroblasts) after adding 8 ug/mL polybrene (Sigma). Medium was changed 6 hours after and cells were sorted based on the positivity of GFP after one passage.

### Cell stimulation

Patient’s and control SV40-fibroblasts were stimulated with 100 ng/mL LTα1β2 (R&D Systems 8884-LY-025/CF) or 10 ng/mL Tweak (PeproTech 310-06) to stimulate the non-canonical pathway of NF-κB and 10 ng/mL TNFα (R&D Systems, #210-TA-020) to stimulate the canonical NF-κB pathway. Poly(I:C) (HMW) from Invivogen was used in the bulk RNA sequencing experiment.

### Real time quantitative PCR

Total RNA was isolated using the RNeasy Mini Kit (Qiagen), and cDNA was prepared using the Quantitect Reverse Transcription Kit (Qiagen) after depletion of genomic DNA. SYBR Select Master Mix was used to perform Real-time quantitative PCR using the LightCycler VIIA7 System (Roche), following manufacturer instruction. The following primers were used to quantify *VCAM1* Fw 5’ GAT TCT GTG CCC ACA GTA AGG C 3’ Rv 5’ TGG TCA CAG AGC CAC CTT CTT G 3’; *TNFAIP3* Fw 5’ TCC TCA GGC TTT GTA TTT GAG C 3’ Rv 5’ TGT GTA TCG GTG CAT GGT TTT A 3’; *CHUK* Fw 5’ CTC GGA AAC CAG CCT CTC AAT G 3’ Rv 5’ GAT AAA CTT CTG GAA GCA AAT GGC 3’. Quantification was normalized to *GAPDH*. RNA expression was analyzed using the ΔΔCt method. All primers were checked for linearity by serial dilution of a control cDNA. A R^2^ > 0.99 was obtained for each set of primers and deltaCT between 10-fold dilution was always 3.2 Ct.

### Bulk RNA sequencing

RNA was extracted from SV40-fibroblasts stimulated or not with TNFα 10 ng/mL or poly I:C 10 ug/mL (High Molecular Weight, Invivogen) for 6 hours. Extraction was performed using the Zymo Quick-RNA 96™ Kit (R1052) following manufacturer instructions with the DNAse step. RNA integrity and concentration were assessed by capillary electrophoresis using Fragment Analyzer (Agilent Technologies). RNaseq libraries were prepared starting from 500 ng of total RNA using the Universal Plus mRNA-Seq kit (Nugen) as recommended by the manufacturer. The oriented cDNA produced from the poly-A+ fraction was sequenced on a NovaSeq6000 from Illumina (Paired-End reads 100 bases + 100 bases). A total of ∼50 million of passing-filters paired-end reads was produced per library. Paired-end RNA-seq reads were aligned to the human Ensembl genome GRCh38.91 using STAR (v2.7.5a) and counted using featureCounts from the Subread R package. The raw count matrix was analyzed using DESeq2 (version 21.38.3). Pre-filtering to keep only rows that have a count of at least 10 for a minimal number of samples have been performed. Differential expression analysis was performed using the “DESeq” function with default parameters. For visualization and clustering, the data was normalized using the ‘variant stabilizing transformation’ method implemented in the “vst” function. Plots were generated using ggplot2 (version 3.4.4), and pheatmap (version 1.0.12).

Pathway activity inference analysis of bulk RNA sequencing data was performed using decoupleR package (version 2.5.3), using default parameters. Visualization of results was performed using ggplot2 (version 3.4.4). Ingenuity Pathway Analysis (IPA, QIAGEN Inc.) was also used to perform pathway activity inference and decipher molecules predicted to be inhibited or activated. All results of the differentially expressed genes between groups, the decoupleR and IPA analysis are available as a supplementary table 2.

### Whole cell lysates

Whole cell lysates from SV40-fibroblasts, activated T-cells and HEK293T cells were prepared by using RIPA buffer (ThermoFischer), supplemented with protease inhibitor cocktail (Thermo Scientific, #87785) and phosphatase inhibitors cocktails 2 and 3 (Sigma P5726-1ML, Sigma P0044-1ML) following manufacturer instructions (complete RIPA buffer). For adherent cells, culture wells were washed 3 times with cold PBS and complete RIPA buffer was added in the well. After 5 min, all cells were lysed in the well, the cell lysate was transferred in aliquots and sonicated using the Bioruptor sonication device previously cooled at 4°C (Diagenode #B01060010) with 10 cycles of 30 seconds on, 30 seconds off. Cell lysates were centrifugated full speed for 5 min on a microcentrifuge at 4°C and supernatants (protein extracts) were collected. Proteins were quantified using the Pierce BCA Protein Assay kit (Thermo Scientific, #23225) just before use. Protein extracts were used immediately or frozen at –20°C for short term conservation and –80°C for long term conservation.

### Cyto-nuclear extractions

Cells were harvested, washed 3 times with cold PBS and centrifuged at 600g for 5 min. The NE-PER protein extraction system was used to extract cytoplasmic and nuclear protein fractions (Thermo Scientific, #78833), following manufacturer instructions. Briefly, after centrifugation, the CER 1 buffer supplemented with protease inhibitor cocktail (Thermo Scientific, #87785) and phosphatase inhibitors cocktails 2 and 3 (Sigma P5726-1ML, Sigma P0044-1ML) was added to the cell pellet, vortexed and placed on ice for 10 min. After 10 min, CER 2 was added for 1 min and cells were centrifugated at > 14000g on a microcentrifuge a 4°C. The supernatant was collected (cytoplasmic fraction) and freezed. The remaining pellet of the aliquot was washed two times with cold PBS and NER buffer supplemented as previously described was added to the remaining pellet. During 40 min, the aliquots were sonicated for 10 min and vortexed every 10 min. The nucleus lysate was centrifugated at maximum speed (>14 000 g) at 4°C for 10 min and the supernatant was collected and freezed (nuclear fraction).

### Western blots

Samples were denatured and reduced with Bolt LDS Sample buffer 4X (Thermo Fisher) and Bolt sample reducing agent 10X (Thermo Fisher). The migration was carried out on 8% Bis-Tris gels (Thermo Fisher) at 150V. The transfer was done on PVDF membranes (Thermo Fisher, #IB24002) with the iBlot 2 dry transfer system (Thermo Fisher, #IB21001) using a the P0 program. The membranes were blocked with 5% bovine serum albumin (BSA) in TBS (Tris Buffer saline) Tween 0.1% for 1 hour at RT (room temperature) with shaking, then incubated with the primary antibody with stirring at 4° C overnight. After 3 washes with TBS Tween 0.1% (TBST), the anti-rabbit or anti-mouse secondary antibody coupled to HRP (Horse Radish Peroxidase) was incubated for 1 hour at RT with stirring at a dilution of 1:10,000 in 5% BSA TBST. After 3 washes with TBST, the membrane was incubated for 5 min, at room temperature (RT), in the dark, with the HRP substrate, contained in the commercial solution SuperSignal West Pico PLUS Chemiluminescent Substrate (Thermo Fisher, # 34580), then developed on a ChemiDoc (Biorad) or Fusion FX (Vilber).

### EMSA experiments

Control and patient fibroblasts were stimulated with 100 ng/mL LTα1β2 (R&D Systems 8884-LY-025/CF) at the indicated time points. Nuclear extracts were prepared and analyzed for DNA-binding activity by using the HIV-LTR tandem kB oligonucleotide as a kB probe.

### Proliferation assay of activated T cells

Day 10 of culture activated T cells were starved of IL-2 for 72h (after 3 washes in PBS and cultivated in complete Panserin without IL-2), washed, incubated with CellTrace violet reagent (Invitrogen) for 817min at 3717°C in the dark, and washed twice more. A total of 2 ×10^5^ cells were seeded into 96-well plates and subjected to different stimuli (plate-bound anti-CD3 antibody (Invitrogen), soluble anti-CD3/CD28 beads (Thermofisher, #11141D). The cells were cultured for 4 days, washed with PBS, and stained with anti-CD3 (PE-Cy7, Biolegend), CD4 (PerCP5.5, Biolegend), CD8 (APC, Biolegend), CD25 (BV650, Biolegend), and CD69 (PE, ThermoFischer) antibodies prior to flow cytometry measurement on a Novocyte (Agilent). Analysis was carried out on the online software OMIQ (OMIQ by Dotmatics) as well as data visualization.

### Detection of autoantibodies using a luciferase reporter assay

The blocking activity of anti-IFN-α2, anti-IFN-ω and anti-IFNβ auto-antibodies (auto-Abs) was determined with a reporter luciferase assay. Briefly, HEK293T cells were transfected with a plasmid containing the firefly luciferase gene under the control of the human ISRE promoter in the pGL4.45 backbone, and a plasmid constitutively expressing Renilla luciferase for normalization (pRL-SV40). Cells were transfected in the presence of the X-tremeGene 9 transfection reagent (Sigma Aldrich, ref. number 6365779001) for 24 hours. Cells in Dulbecco’s modified Eagle medium (DMEM, Thermo Fisher Scientific) supplemented with 2% fetal calf serum (FCS) and 10% healthy control or patient serum/plasma were either left unstimulated or were stimulated with IFN-α2 (Miltenyi Biotec, ref. number 130-108-984) or IFN-ω (Merck, ref. number SRP3061) at 10 ng/mL or 100 pg/mL, or with IFN-β (Miltenyi Biotech, ref. number: 130-107-888) at 10 ng/mL for 16 hours at 37°C. Each sample was tested once for each cytokine and dose, in at least two independent experiments. Finally, cells were lysed for 20 minutes at room temperature and luciferase levels were measured with the Dual Luciferase Reporter 1000 assay system (Promega, ref. number E1980), according to the manufacturer’s protocol. Luminescence intensity was measured with a VICTOR X Multilabel Plate Reader (PerkinElmer Life Sciences, USA). Firefly luciferase activity values were normalized against Renilla luciferase activity values. These values were then normalized against the plasma used in non-stimulated conditions. Samples were considered to be neutralizing if the luciferase activity signal, normalized against non-stimulated conditions, was below 5.

### CyTOF staining

PBMCs from the patient and controls were analyzed using CyTOF (Maxpar® Direct™ Immune Profiling System Fluidigm, Les Ulus, France) including a 30-marker antibody panel (Fluidigm, ref. #201325). 3×106 PBMCs resuspended in 300μl of MaxPar Cell Staining Buffer were incubated for 10 minutes at room temperature after addition of 5μl of Human TruStain FcX (Biolegend Europ, Netherland). Then the 300 μl were directly added to the dry antibody cocktail containing 30 antibodies, for 30 minutes. All samples were washed three times in MaxPar Cell Staining Buffer and then fixed in 1.6% paraformaldehyde (Sigma-Aldrich, France) for 10 minutes. After centrifugation, the PBMCs were incubated overnight in Fix and Perm Buffer with 1:1000 of iridium intercalator (pentamethylcyclopentadienyl-Ir (III)-dipyridophenazine, Fluidigm, Les Ulis, France) and frozen at –80°C before acquisition.

### CyTOF acquisition

After thawing, cells were washed and resuspended at a concentration of 1 million cells per mL in Maxpar Cell Acquisition Solution, a high-ionic-strength solution, and mixed with 10% of EQ Beads immediately before acquisition. Samples were acquired on the Helios mass cytometer and CyTOF software version 6.7.1014 (Fluidigm, Inc Canada) at the “Plateforme de Cytométrie de la Pitié-Salpétriêre (CyPS)”. An average of 500,000 events were acquired per sample. Dual count calibration, noise reduction, cell length threshold between 10 and 150 pushes, and a lower convolution threshold equal to 10 were applied during acquisition. Mass cytometry standard files produced by the HELIOS were normalized using CyTOF Software v. 6.7.1014. Four parameters (center, offset, residual, and width) are used for data cleaning to resolve ion fusion events (doublets) from single events from the Gaussian distribution generated by each event. After data cleaning, the program produces new FCS files consisting of only intact live singlet cells.

### CyTOF analysis

FCS files containing intact live singlet cells were uploaded in R version 4.0.3 using the flowCore package. All files were concatenated in a SingleCellExperiment (SCE) with the SingleCellExperiment package. After quality control (number of cells per sample, expression pattern of all markers across samples), all cells were submitted to clustering using the cluster function (FlowSOM and ConsensusMetaClustering) following the recommendation from the CATALYST packages (1). For clustering, the k parameter was set to 60 to detect small immune populations. Clusters were then identified based on their expression of type markers according to previous knowledge of immune cell phenotype and following manufacturer instruction from Fluidigm Maxpar Immune Profiling. Clusters expressing the same cell type markers were merged into one immune cell population. The proportion of clusters among all intact viable cells was then defined using the appropriate function from CATALYST and compared between groups using the Mann-Whitney Wilcoxon test in R. For visualization purposes, dimensionality reduction in UMAPs was performed using the runDR function from CATALYST with neighbors set to 15 and minimum distance to 0.4. All other visualizations were performed using in-house and the ggplot2 function. Subclustering of memory T cells was performed on all memory CD4+ T cells (excluding Tregs) based on the markers CXCR3, CXCR5, CCR6 and CCR4 (defined as type markers in a new panel file). Subclustering of B cells were performed based on the following markers: CD19, CD11c, CD27, CXCR5, CD38, CD20, HLA-DR, IgD.

### Statistical analysis and data representation

All statistical analyses were performed with R version 4.2.3. As indicated in the Figures, results were analyzed with appropriate parametric or non-parametric statistical tests. For all analyses, the threshold for statistical significance was set to p<0.05. Data visualization was made either in R with ggplot2 and ggpubr packages or GraphPad Prism.

---

**Supplementary Figure 1.**
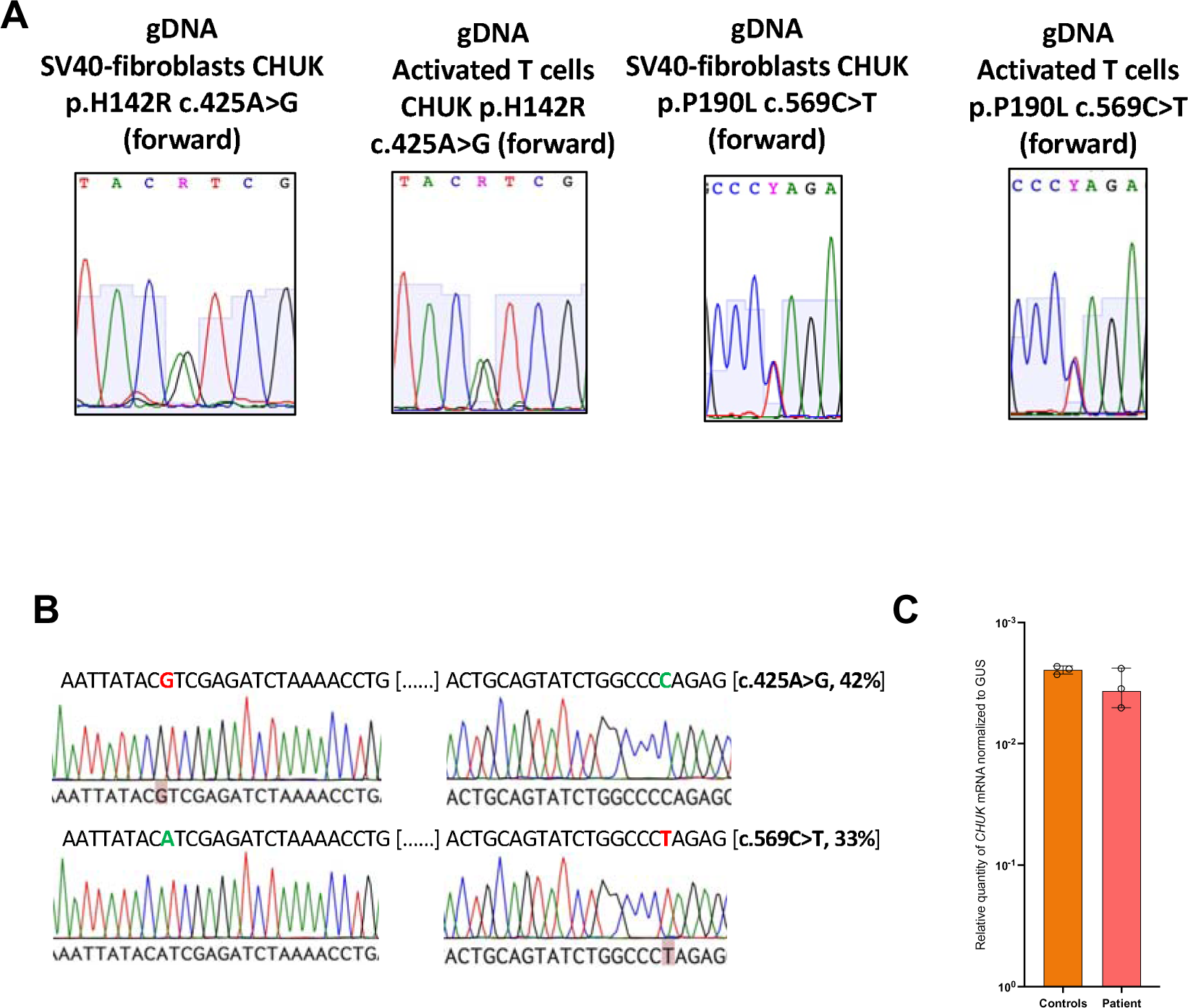
Sanger sequencing on different cell types from the patient and healthy controls. **A**. Sanger sequencing of the patient mutation on genomic DNA **B.** Example of TOPO-TA cloning results from RT-PCR products encompassing the two mutations of the patient; Frequency of the corresponding sequence is written on the right. **C**. Relative quantity of *CHUK* mRNA in SV40-fibroblast from patient and controls assessed by RTqPCR

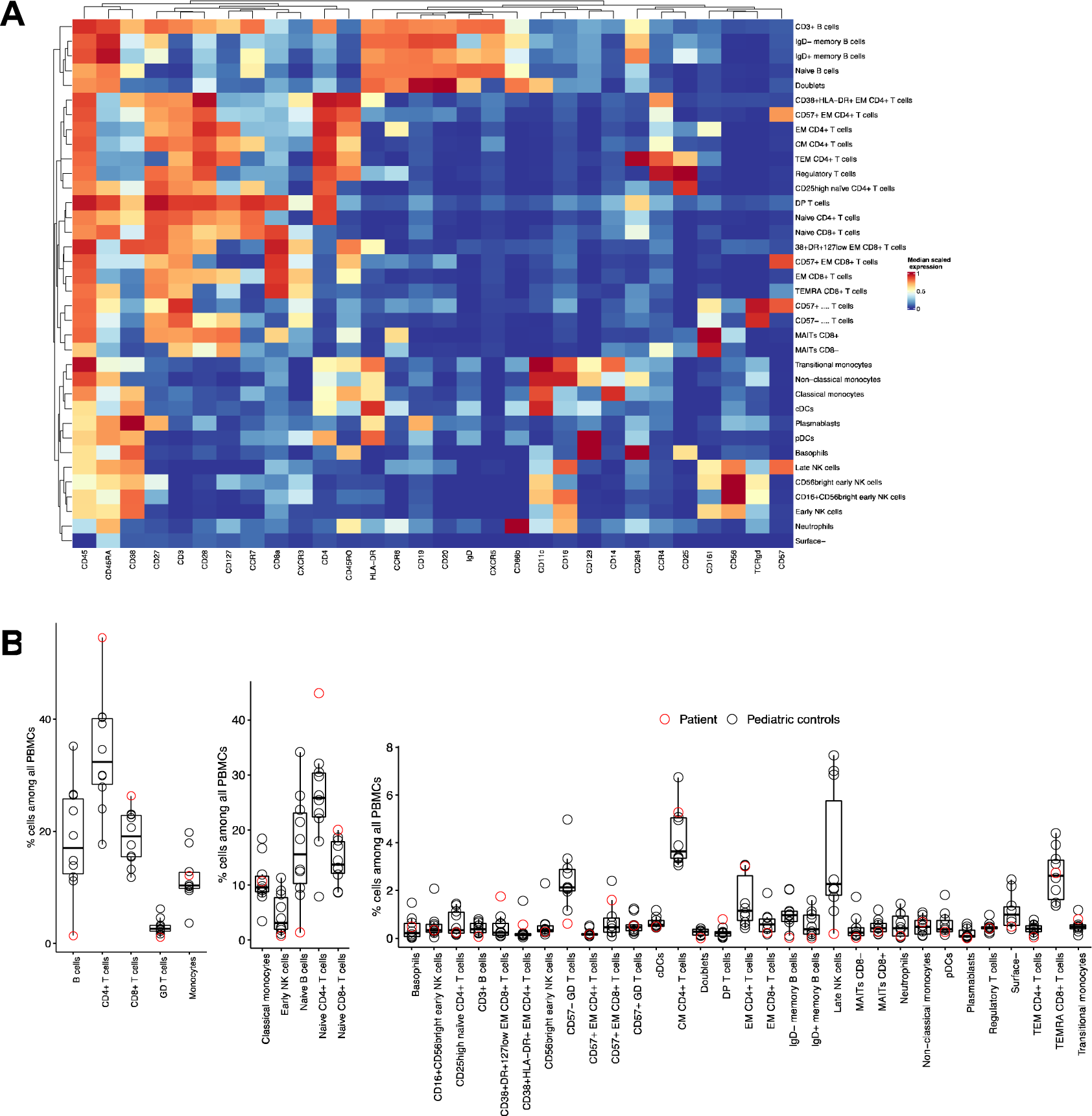
Supplementary Figure 2.

**Supplementary Figure 3.**
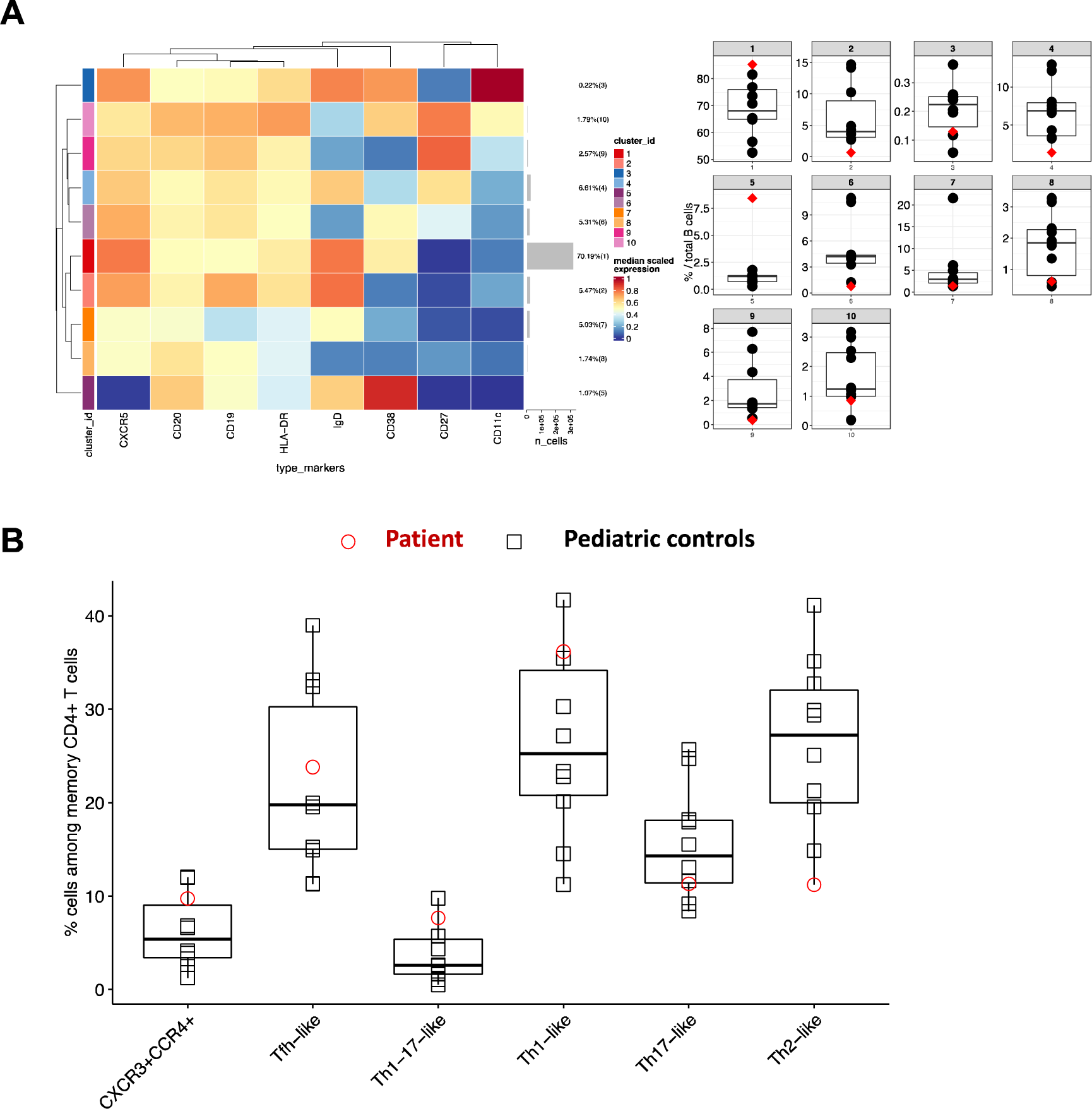
CD4+ T cells were subjected to reclustering to identify T-helper cell subsets. The relative abundance of each identified clusters is shown (patient in red, controls in black).

**Supplementary Figure 4.**
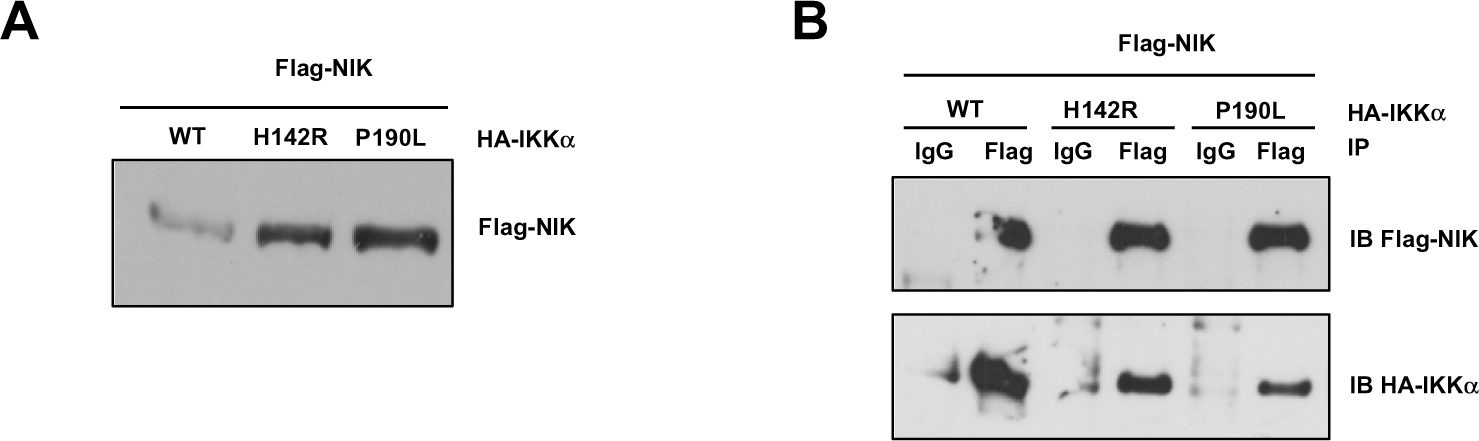
Co-immunoprecipitation between IKKα and NIK. **A**. Whole cell lysate of HEK293T cells co-transfected with Flag-NIK and HA-IKKa wt or mutants (as indicated) were subjected to western-blot. Immunoblot of Flag-NIK. **B**. Immunoprecipitation of Flag-NIK after transfection of HEK2093T cells with Flag-NIK and HA-IKKα wt or mutants (as indicated). Immunoblot of Flag-NIK and HA-IKKα.

**Supplementary Figure 5.**
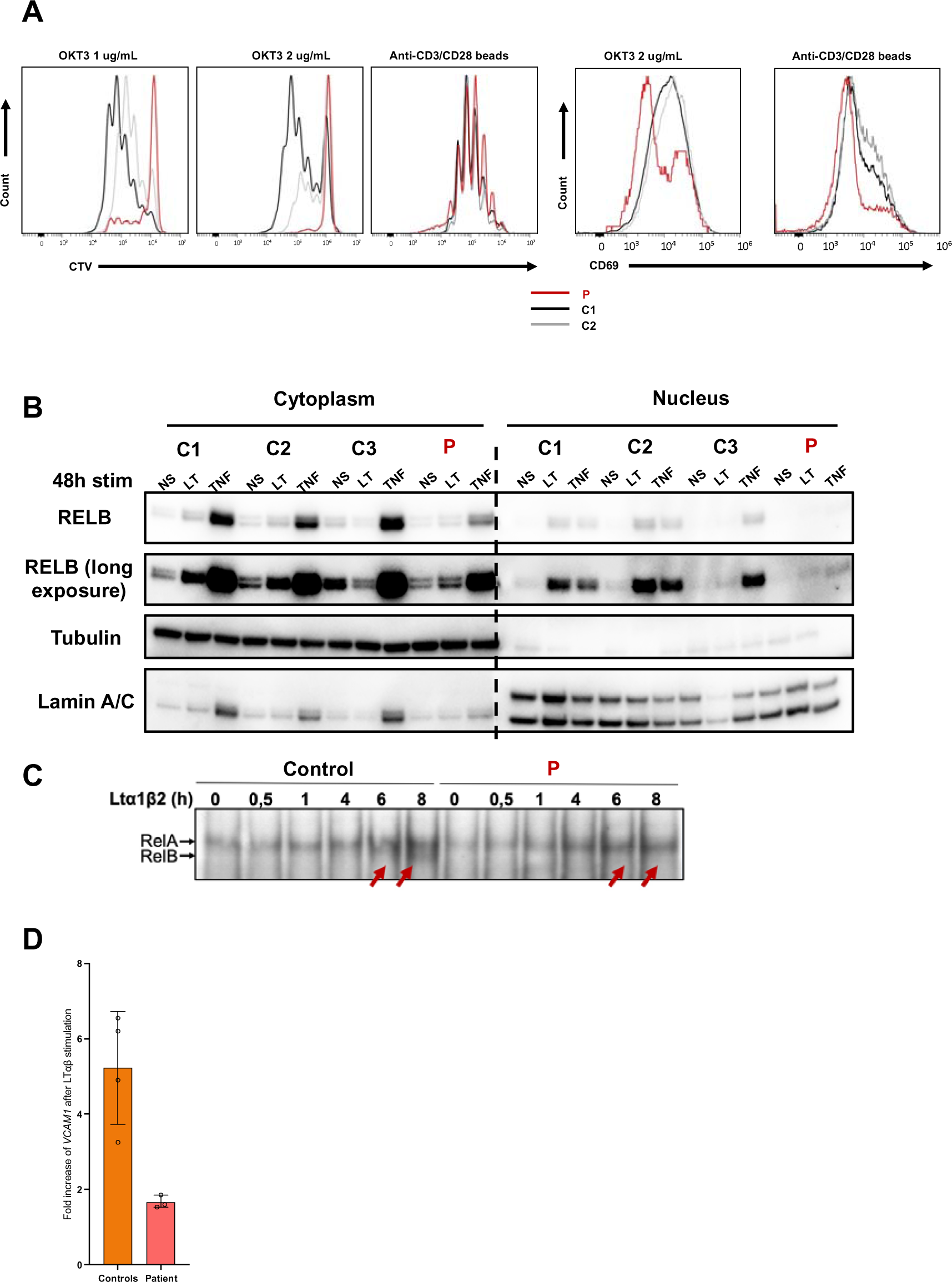
Functional impact of the variant on non-canonical and canonical NF-κB pathways. **A**. Dilution histogram of Cell Trace Violet dye (CTV) after stimulation of activated T-cells with OKT3 at 1 or 2 ug/mL for 4 days or anti-CD3/CD28 beads for 4 days. Histogram of CD69 activation marker upregulation on activated T cells after 4 days of stimulation with either OKT3 or anti-CD3/CD28 beads. Patient is in red, controls in gray. **B**. Western-blot of cytoplasmic and nuclear extracts from SV40-fibroblasts after 48h of LTαβ (LT) or TNFα (TNF) stimulation. RELB was revealed using a specific antibody to assess for RELB production and nuclear translocation. Tubulin was used a loading control of cytoplasmic extract while Lamin A/C was used for nuclear extracts. The patient is indicated in red, the controls are C1, γC2 and C3. **C**. Electromobility shift assay on nuclear extracts from healthy control and patient SV40-fibroblasts after different time points of LTα1β2 stimulation showing the absence of RELB binding (red arrows) in patient as compared to controls (lower thin band) and the normal binding of RELA (upper band). **D**. Relative *VCAM1* mRNA expression after 24h of LTαβ relative to non-stimulated condition (left panel). These experiments have been performed on healthy controls and patient SV40-fibroblasts.

**Supplementary Figure 6.**
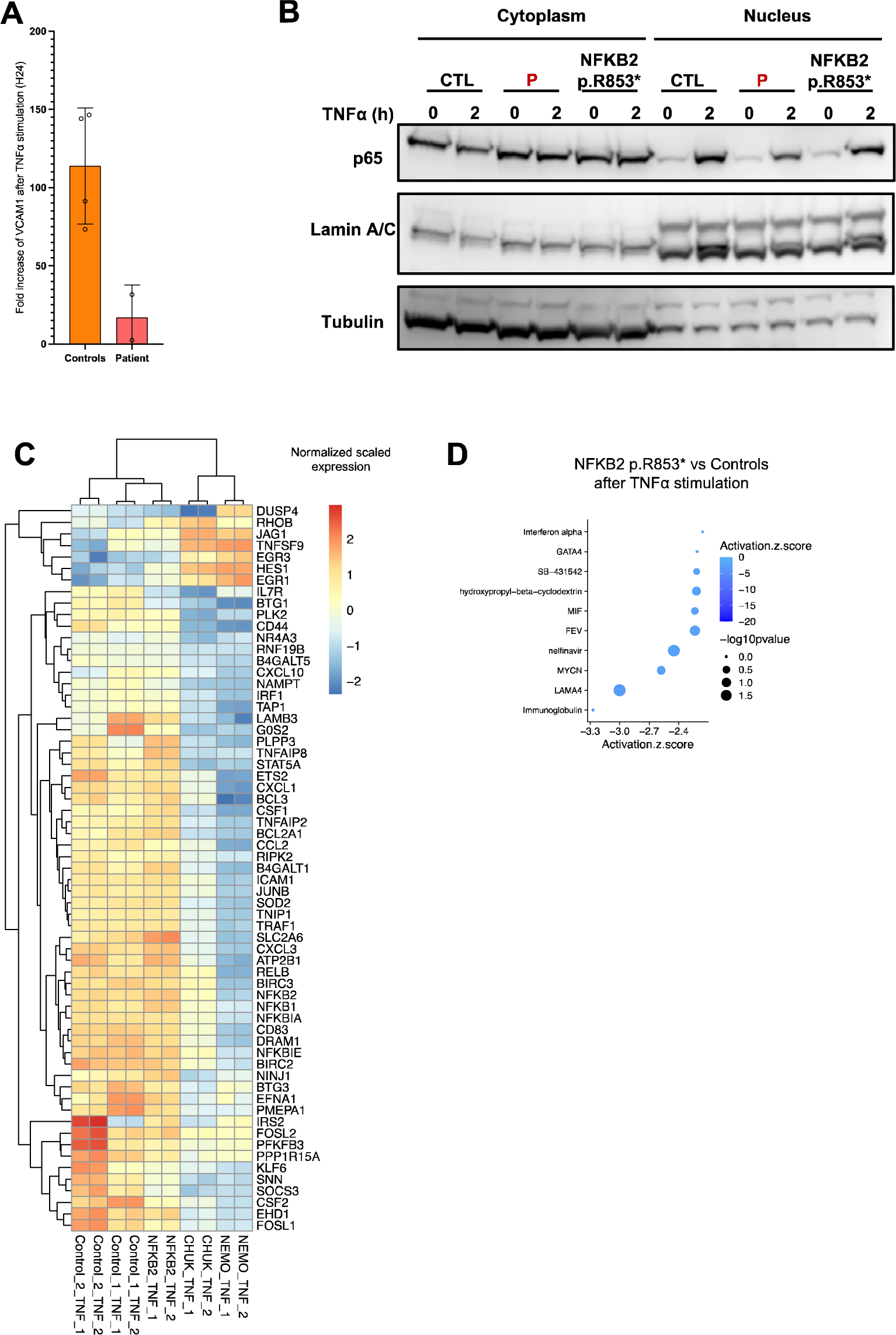
Bulk RNA sequencing of SV40-fibroblasts stimulated with TNFα. **A**. Relative *VCAM1* mRNA expression after 24h of TNFα relative to non-stimulated condition (left panel). **B**. Western-blot of cytoplasmic and nuclear protein extracts from SV40 fibroblasts activated with TNFα for 2 hours assessing the translocation of p65/RELA in the nucleus in patient’ cells (P), a patient with a known p52 LOF/p100 IκBδ GOF heterozygous mutation in *NFKB2* (NFKB2 p.R853*). Tubulin was used a loading control of cytoplasmic extract while Lamin A/C was used for nuclear extracts. LOF: loss-of-function, GOF:gain-of-function **C.** Heatmap showing the normalized and scaled expression (vst) of genes significantly upregulated in control SV40-fibroblasts after stimulation with TNFα 10 ng/mL for 6 hours and significantly downregulated in SV40-fibroblasts from the patient compared to the controls after TNFα stimulation. Each column represents a patient or a control. RNA sequencing has been performed in biological duplicates for each control or patient (as indicated by _1 or _2 at the end of each column name). **D.** Z-score of the different “molecules” predicted to be inhibited (top 10 based on the z-score) based on Ingenuity Pathway Analysis (IPA, QIAGEN Inc) for the NFKB2 p.R853* patient. A negative score indicate that the molecule is predicted to be inhibited based on the differentially expressed genes that have been processed.

**Supplementary Figure 7.**
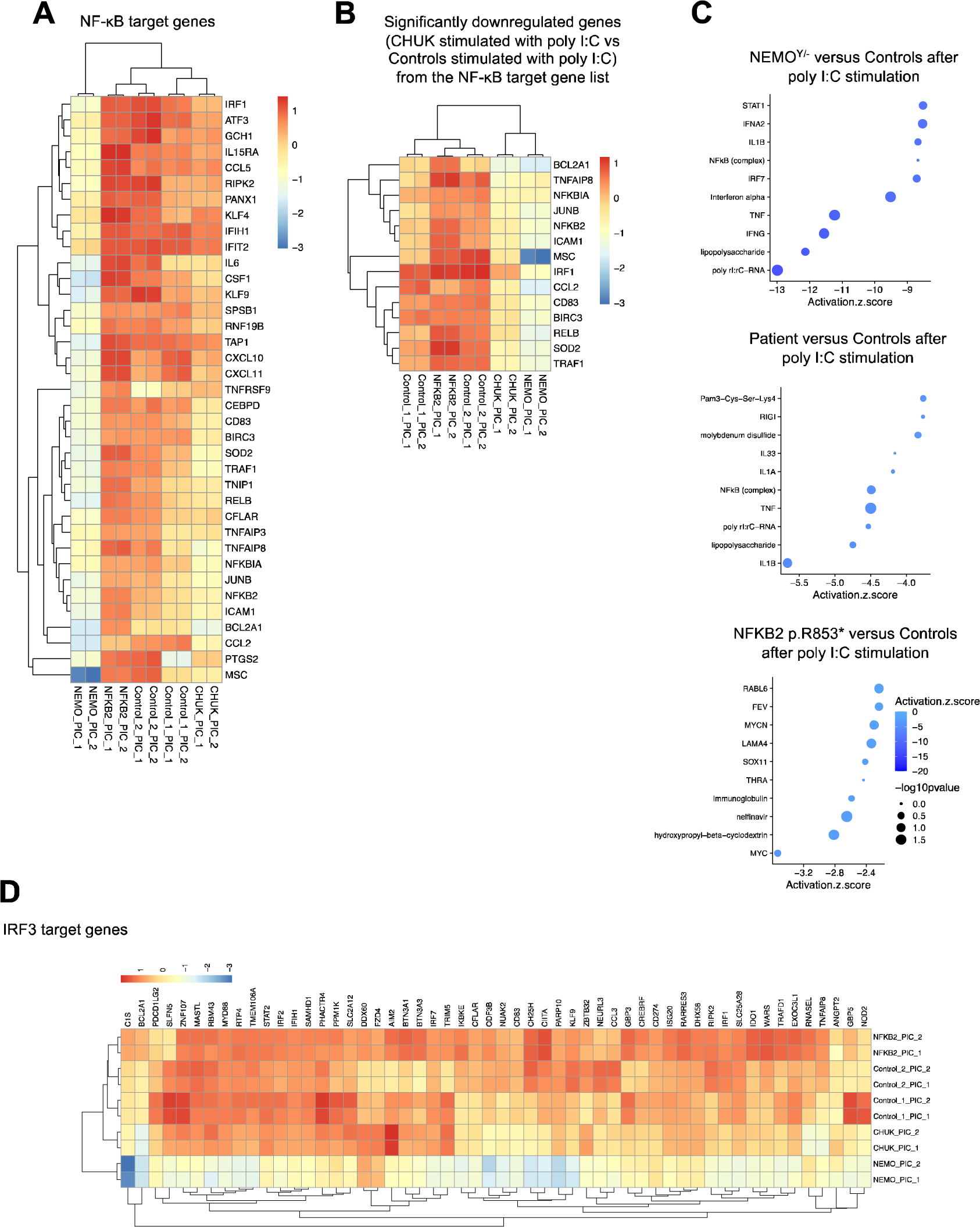
Bulk RNA sequencing of SV40-fibroblasts stimulated with poly I:C. **A**. Heatmap showing the normalized and scaled expression (vst) of NF-κB target genes significantly upregulated in controls after stimulation with poly I:C 10 ug/mL for 6 hours. Each column represents a patient or a control. RNA sequencing has been performed in biological duplicates for each control or patient (as indicated by _1 or _2 at the end of each column name). **B**. Heatmap showing the normalized and scaled expression (vst) of genes significantly upregulated in controls after stimulation with poly I:C 10 ug/mL for 6 hours represented in the MSIgDB database TNFa via NF-κB signaling (curated geneset of NF-κB target genes). **C**. Z-score of the different “molecules” predicted to be inhibited (top 10 based on the z-score) based on Ingenuity Pathway Analysis (IPA, QIAGEN Inc) for indicated comparisons. A negative score indicate that the molecule is predicted to be inhibited based on the differentially expressed genes that have been processed in IPA. **D**. Heatmap showing the normalized and scaled expression (vst) of IRF3 target genes significantly upregulated in controls after stimulation with poly I:C 10 ug/mL for 6 hours.

**Supplementary Table 1. Routine immune phenotype of the patient’ whole blood**.

**Supplementary Table 2. Differentially expressed genes from the bulk RNA sequencing analysis.**

